# Dietary Fatty Acids and Epigenetic Aging in US Adults: Results from the National Health and Nutrition Examination Survey

**DOI:** 10.1101/2025.07.21.25331937

**Authors:** Anne Bozack, Dennis Khodasevich, Jamaji C. Nwanaji-Enwerem, Nicole Gladish, Hanyang Shen, Saher Daredia, Belinda L. Needham, David H. Rehkopf, Marta Guasch-Ferre, Andres Cardenas

## Abstract

Fatty acids are involved in disease risk and aging processes. In the US National Health and Nutrition Examination Survey (1999-2002), we tested for associations of total, saturated (SFA), monounsaturated (MUFA), polyunsaturated (PUFA), and subtypes of dietary fatty acids with DNA methylation-based aging biomarkers, adjusting for age, BMI, total energy intake, and sociodemographic and behavioral factors (N=2,260). Higher SFA and MUFA were associated with greater GrimAge2, an aging biomarker of mortality; PUFA was associated with lower Horvath1, Hannum, and PhenoAge (*p*<0.05). Omega-3 and the PUFA:SFA ratio were negatively associated with Horvath1, Hannum, Vidal-Bralo, and PhenoAge. Notably, a one-unit increase in PUFA:SFA was associated with 1.05 years lower PhenoAge (95% CI=-1.87, −0.22). There were consistent trends of positive associations of SFA subtypes and negative associations of PUFA subtypes with epigenetic aging; associations of MUFA subtypes varied. Future studies, including randomized controlled trials, are needed to investigate causality and downstream clinical outcomes.

## Introduction

As the global population undergoes a demographic shift to a greater proportion of older adults, it is crucial to understand factors that contribute to healthy aging. Beyond genetic determinants, health behaviors throughout life, including physical activity, socializing, and dietary patterns, can help to delay the onset of age-related decline or morbidities and maintain functional longevity.^1^ In particular, nutrition may contribute to healthy aging^2^ or, conversely, increased risk of disease, by impacting pathways related to inflammation, oxidative stress, DNA repair, regenerative potential, and insulin sensitivity.^3^ The purpose of this study is to examine associations of fatty acid intake with DNA methylation-based aging biomarkers.

As one of the major macronutrient groups, lipids, or fatty acids, are necessary to support health through critical biological roles as energy sources, constituents of cell membranes, regulators of signaling pathways and gene expression, and building blocks of hormones.^4^ Fatty acids are classified based on their hydrocarbon chain length and structure and are grouped into saturated (SFAs), monounsaturated (MUFAs), and polyunsaturated fatty acids (PUFAs). SFAs upregulate the synthesis of cholesterol, triglycerides, and phospholipids, and increase the expression of genes involved in inflammatory pathways and insulin sensitivity.^4^ PUFAs include the essential fatty acids omega-6 (ω-6) and omega-3 (ω-3).

Omega-6 fatty acids are major constituents of cell membranes and can lower low-density lipoprotein (LDL) cholesterol levels though the downregulation of biosynthesis.^5^ Omega-3 fatty acid subtypes are abundant in the cell membranes of the brain and eye and influence membrane protein signal transduction.^4^ Omega-3 fatty acids in cell membranes are also involved in controlling inflammation, blood clotting, and constriction of blood vessels.

Recent advancements in the field of epigenetics have enabled the study of age-related health risks and the healthspan through proxies, or biomarkers, of biological aging. Epigenetic clocks, calculated from weighted combinations of DNA methylation levels at age-associated loci, have been developed to estimate chronological age (i.e., first generation clocks), phenotypic age (i.e., second generation clocks), and pace of aging.^6^ Epigenetic clocks are valuable in their ability to capture broad changes in physiological pathways, functional decline, chronic diseases, and damage on the cellular and molecular levels. The difference between chronological age and epigenetic age, known as epigenetic age deviation or epigenetic age acceleration, is associated with morbidities and all-cause mortality.^7,8^ Second-generation clocks, in particular, are strong predictors of cardiometabolic diseases, cancer, and time to death.^9–11^ Leveraging epigenetic clocks may allow for the identification of factors that increase the risk of age-related decline and disease, as well as factors that promote healthy aging and extend functional longevity. Moreover, epigenetic clocks hold promise as predictive and prognostic biomarkers in clinical settings and to inform individual-level diet and behavioral interventions.

There is increasing evidence linking diet quality and fatty acids to disease risk and aging processes.^4,12,13^ However, further research is needed to understand how intake of fatty acids and fatty acid subtypes is related to epigenetic aging. We investigated cross-sectional associations of fatty acid dietary intake and epigenetic aging biomarkers in the United States (US) National Health and Nutrition Examination Survey (NHANES), a representative sample of the US adult population. In particular, we tested for associations of total fatty acids, SFAs, MUFAs, PUFAs, omega-6, omega-3, the ratio of PUFA:SFA (P:S), and 19 fatty acid subtypes. We hypothesized that higher intake of SFAs would be associated with greater epigenetic aging and that higher intake of PUFAs and the P:S ratio would be associated with lower epigenetic aging.

## Results

We summarize DNA methylation biomarkers of aging measured in NHANES, their classification, and their characteristics in **Table 1**. Characteristics of participants with DNA methylation, dietary, and covariate data included in our primary analyses of complete cases (N = 1,771) are shown in **Table 2**, and characteristics of participants included in our imputation analyses (N = 2,220) are shown in **Supplemental Table S1**. Participants in our primary analyses had a mean (SD) age of 64.8 (9.3) years, and half were male (n = 952; 53.8%). Fatty acid intake was estimated by NHANES from 24-hour dietary recall questionnaires. The mean (SD) daily intake in g of total fatty acids, SFAs, MUFAs, PUFAs, omega-6, and omega-3 was 69.2 (37.3), 21.8 (13.3), 25.7 (15.0), 14.6 (9.3), 13.0 (8.4), and 1.4 (1.0), respectively, and the mean (SD) of the PUFA:SFA (P:S) ratio was 0.77 (0.45). Participants reported consuming an average of 1,860 (764) kcal/day, and the mean (SD) percentage of total energy from total fatty acids, SFAs, MUFAs, and PUFAs was 32.9% (9.3%), 10.3% (3.9%), 12.1% (4.1%), and 7.0% (3.2%), respectively. Characteristics among participants included in imputation analyses were similar (**Supplemental Table S1**). Means and SDs of fatty acid subtypes are shown in **Supplemental Table S2**.

**Table 1:** Summary and description of epigenetic clocks and biomarkers included in the study.

| Clock or biomarker | Tissue | Prediction | # of training samples | # of CpGs |
| --- | --- | --- | --- | --- |
| **First generation – chronological age predictors** |  |  |  |  |
| Horvath1 (pan-tissue)<sup>37</sup> | 51 human tissues | Chronological age | 3,931 | 353 |
| Horvath2 (skin & blood)<sup>50</sup> | Blood, cord blood, skin, fibroblast, epithelium, buccal | Chronological age | 869 | 391 |
| Hannum<sup>38</sup> | Blood | Chronological age | 656 | 71 |
| Lin<sup>51</sup> | Blood | Chronological age | 656 | 99 |
| Zhang<sup>41</sup> | Blood and saliva | Chronological age | 13,566 | 514 |
| Vidal-Bralo<sup>42</sup> | Blood | Chronological age | 390 | 8 |
| **Second generation – aging phenotype predictors** |  |  |  |  |
| PhenoAge<sup>9</sup> | Blood | Phenotypic age | 456 | 531 |
| GrimAge2<sup>11</sup> | Blood | Lifespan | 1,833 | 1,030 |
| DNAm telomere length (DNAmTL)<sup>52</sup> | Blood | Telomere length | 2,256 | 140 |
| Epigenetic timer of cancer (epiTOC)<sup>53</sup> | Blood | Cell divisions | 656 | 385 |
| **Third generation – pace of aging predictor** |  |  |  |  |
| Dunedin Pace of Aging Methylation (DunedinPoAm)<sup>36</sup> | Blood | Pace of aging | 810 | 46 |

**Table 2:** Participant characteristics (N = 1,771).

|  | Mean (SD) or n (%) |
| --- | --- |
| Male, n (%) | 952 (53.8%) |
| Age (years) , mean (SD) | 64.8 (9.3) |
| Race and ethnicity, n (%) |  |
| Mexican American | 493 (27.8%) |
| Other Hispanic | 100 (5.6%) |
| Non-Hispanic White | 754 (42.6%) |
| Non-Hispanic Black | 366 (20.7%) |
| Other including Multiracial | 58 (3.3%) |
| BMI (kg/m<sup>2</sup>), mean (SD) | 28.9 (5.9) |
| Smoking status, n (%) |  |
| Never | 780 (44.0%) |
| Former | 713 (40.3%) |
| Current | 278 (15.7%) |
| Alcohol intake, drinks/day, mean (SD) | 0.4 (0.9) |
| Education, n (%) |  |
| Less Than High School | 728 (41.1%) |
| High School Diploma (including GED) | 388 (21.9%) |
| More Than High School | 655 (37.0%) |
| Occupation, n (%) |  |
| Blue-collar and semi-routine | 704 (39.8%) |
| Blue-collar and high skill | 250 (14.1%) |
| White-collar and semi-routine | 323 (18.2%) |
| White-collar and professional | 452 (25.5%) |
| No work as described | 42 (2.4%) |
| PIR, mean (SD) | 2.7 (1.6) |
| Energy (kcal), mean (SD) | 1,860 (764) |
| Total fatty acids (g), mean (SD) | 69.2 (37.3) |
| Percent of total energy, mean (SD) | 32.9 (9.3) |
| Saturated fatty acids (g), mean (SD) | 21.8 (13.3) |
| Percent of total energy, mean (SD) | 10.3 (3.9) |
| Monounsaturated fatty acids (g), mean (SD) | 25.7 (15.0) |
| Percent of total energy, mean (SD) | 12.1 (4.1) |
| Polyunsaturated fatty acids (g), mean (SD) | 14.6 (9.3) |
| Percent of total energy, mean (SD) | 7.0 (3.2) |
| Omega-6 (g), mean (SD) | 13.0 (8.4) |
| Omega-3 (g), mean (SD) | 1.4 (1.0) |
| Polyunsaturated:saturated (P:S) fatty acid ratio, mean (SD) | 0.77 (0.45) |

Survey-weighted correlations of the intake of total fatty acids and fatty acid subtypes are shown in **Supplemental Figures S2** and **S3**. As expected, there were moderate-to-strong correlations between the intake of total fatty acids, SFAs, MUFAs, and PUFAs (*r* = 0.54-0.97; *p* < 0.001). The P:S ratio was negatively correlated with total fatty acids, SFAs, and MUFAs (*r* = −0.06, −0.35, and −0.09, respectively; *p* < 0.05), and positively correlated with PUFAs (*r* = 0.43; *p* < 0.01).

Scatter plots of chronological age and each of the epigenetic aging biomarkers analyzed are shown in **Supplemental Figure S4**. Epigenetic age estimates were strongly correlated with chronological age (*r* range = 0.62-0.91). The Zhang clock was most strongly correlated with chronological age (*r* = 0.91) but overestimated the ages of younger participants while underestimating the ages of older participants. The Vidal-Bralo clock, which is based on 8 CpGs, had the lowest correlation with chronological age (*r* = 0.62). As expected, DNAmTL was negatively correlated (*r* = −0.59), epiTOC was positively correlated (*r* = 0.22), and DunedinPoAm had a weak correlation with chronological age (*r* = 0.06).

### Associations of total fatty acid intakes with epigenetic aging biomarkers

We evaluated the associations of fatty acid intake with epigenetic aging biomarkers using adjusted survey-weighted linear models, adjusting for age, age^2^, sex, race and ethnicity, BMI, education level, occupation, poverty to income ratio, smoking status, alcohol intake, physical activity, and total energy intake. Fatty acid intake variables in g were log_2_ transformed prior to analysis.

Total fatty acid intake was not significantly associated with any epigenetic aging biomarker. However, log_2_ increase, or doubling, of SFA and MUFA intake was associated with 0.42 years (95% CI = 0.02, 0.83) and 0.54 years (95% CI = 0.04, 1.04) greater GrimAge2, respectively (**Figure 1**; effect sizes, 95% confidence intervals (CIs), and p-values are shown in **Supplemental Table S2**). Conversely, a doubling in PUFA intake was associated with 0.68 years lower Horvath1 (95% CI = −1.26, −0.09), 0.60 years lower Hannum (95% CI = −1.15, −0.06), and 0.59 years lower PhenoAge (95% CI = −1.14, −0.03). We did not observe any significant associations of fatty acid intake with DNAmTL, epiTOC, or DunedinPoAm.

**Figure 1:**
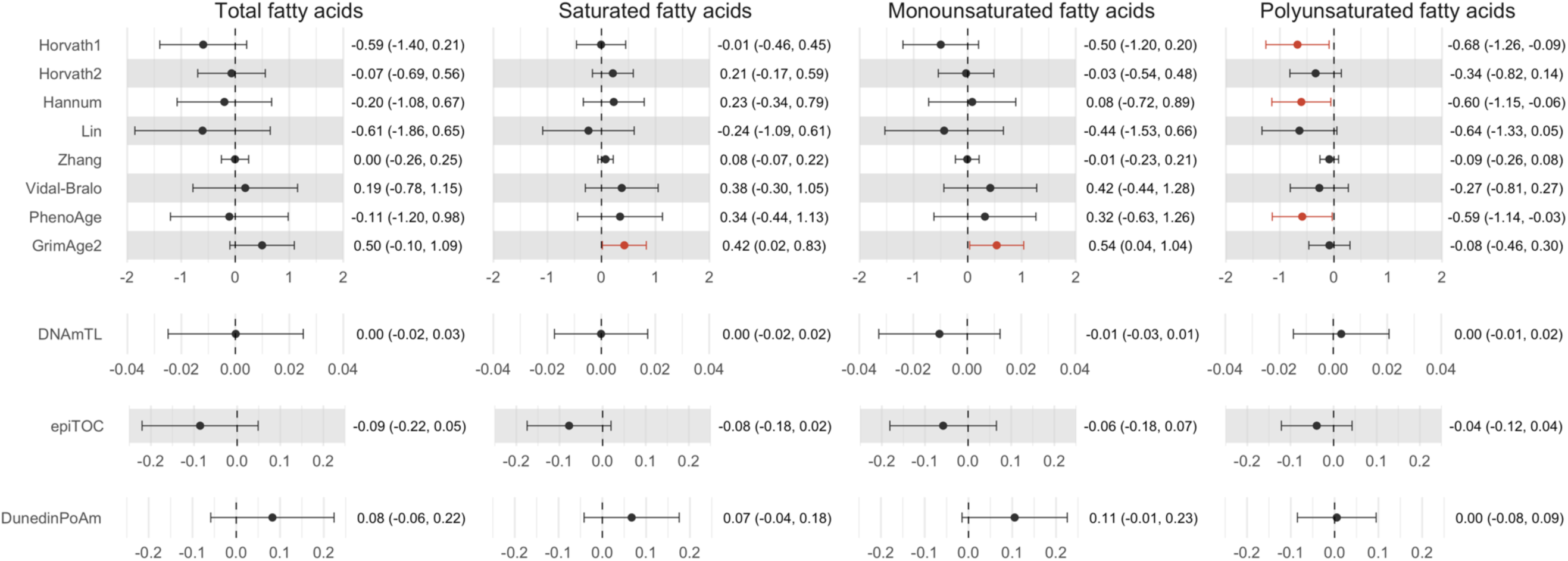
Associations of total, saturated, monounsaturated, and polyunsaturated fatty acid intake with epigenetic aging biomarkers. Effect estimates (95% confidence intervals (CIs)) are shown for a log_2_ increase, or doubling, of fatty acid intake. Units are in years for the Horvath1, Horvath2, Hannum, Lin, Zhang, Vidal-Bralo, PhenoAge, and GrimAge2 clocks; kilobases for DNAmTL, and standard deviations for epiTOC and DunedinPoAm. Results are from weighted generalized linear regression models adjusted for age, age^2^, sex, race and ethnicity, BMI, education level, occupation, poverty to income ratio, smoking status, alcohol intake, physical activity, and total energy intake. Significant associations (*p* < 0.05) are plotted in red.

### Associations of omega-6 and omega-3 with epigenetic aging biomarkers

Across most first- and second-generation epigenetic clocks, there was a trend toward negative associations of omega-6 and omega-3 intake with epigenetic aging (**Figure 2** and **Supplemental Table S2**). Intake of omega-6 was significantly associated with lower epigenetic aging as measured by the Horvath1 and Hannum clocks (*p* < 0.05), while omega-3 intake was significantly associated with lower epigenetic aging as measured by Horvath1, Hannum, Vidal-Bralo, and PhenoAge clocks. The greatest effect size was observed for omega-3 intake and PhenoAge, with a doubling of omega-3 intake associated with 0.77 years lower PhenoAge (95% CI = −1.33, −0.21).

**Figure 2:**
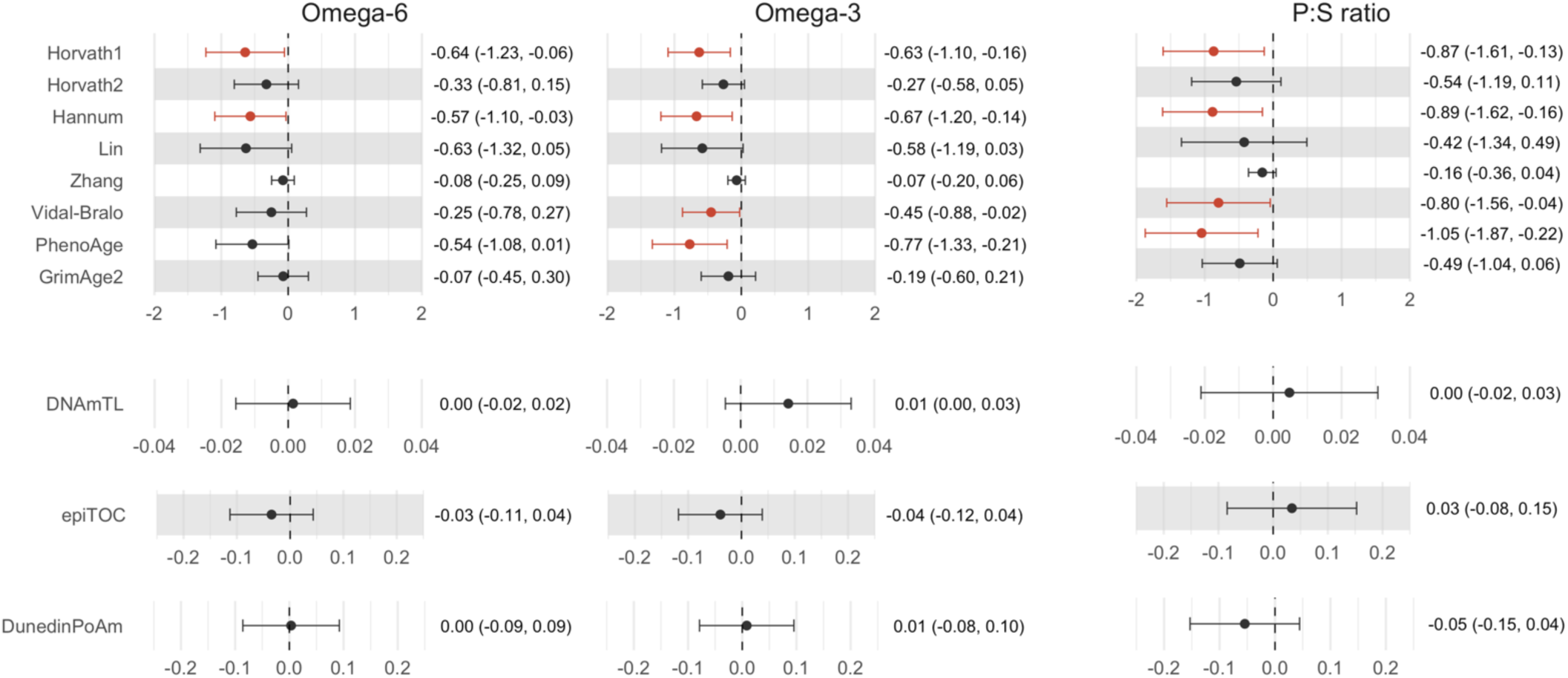
Associations of omega-6 intake, omega-3 intake, and the ratio of polyunsaturated:saturated fatty acids (P:S) with epigenetic aging biomarkers. Effect estimates (95% confidence intervals (CIs)) are shown for a log_2_ increase, or doubling, of omega-6 and omega-3 intake and for a one-unit increase in the P:S ratio, which approximates the interquartile range. Units are in years for the Horvath1, Horvath2, Hannum, Lin, Zhang, Vidal-Bralo, PhenoAge, and GrimAge2 clocks; kilobases for DNAmTL, and standard deviations for epiTOC and DunedinPoAm. Results are from weighted generalized linear regression models adjusted for age, age^2^, sex, race and ethnicity, BMI, education level, occupation, poverty to income ratio, smoking status, alcohol intake, physical activity, and total energy intake. Significant associations (*p* < 0.05) are plotted in red.

Considering that omega-6 and omega-3 were highly correlated (*r* = 0.79, **Supplemental Figure S2**), we conducted analyses simultaneously modeling both variables. The variance inflation factor (VIF) indicated low-to-moderate multicollinearity, with values ranging from 2.9-3.9 and 2.2-3.4 for omega-6 and omega-3, respectively, across the epigenetic aging biomarker models. In jointly adjusted models, omega-6 intake was not significantly associated with any epigenetic aging biomarker (**Supplemental Table S3**). However, the associations of omega-3 intake with Vidal-Bralo and PhenoAge remained significant (*B* (95% CI) = −0.57 years (−1.13, −0.01) and −0.84 years (−1.64, −0.05), respectively).

### Associations of the ratio of polyunsaturated:saturated fatty acids with epigenetic aging biomarkers

The P:S ratio was significantly associated with lower epigenetic aging as measured by three first-generation clocks and PhenoAge (**Figure 2** and **Supplemental Table S2**). A one-unit increase in the P:S ratio was associated with 0.87 years lower Horvath1 (95% CI = −1.61, −0.31), 0.89 years lower Hannum (95% CI = −1.62, −0.16), and 0.80 years lower Vidal-Bralo age (95% CI = −1.56, −0.04). However, like the associations with omega-3 intake, the greatest association was with PhenoAge (*B* (95% CI) = −1.02 years per unit increase (−1.80, −0.25)). A one-unit difference in the P:S ratio approximates the interquartile range (IQR) of 0.92. The associations of the P:S ratio with the Horvath2, Lin, Zhang, and GrimAge2 clocks and with DunedinPoAm were negative but imprecise.

### Associations of fatty acid subtypes with epigenetic aging biomarkers

In analyses of fatty acid subtypes, we found consistent trends in associations across the epigenetic aging biomarkers (**Figure 3** and **Supplemental Tables S3-S5**). Notably, all PUFA subtypes had negative, but imprecise, directions of association with first-generation clocks, PhenoAge, and epiTOC, and positive directions of association with DNAmTL. PUFA 22:6 (DHA) was significantly associated with the greatest number of epigenetic aging biomarkers: Horvath1, Horvath2, Hannum, Zhang, PhenoAge, and DNAmTL. The association of PUFA 18:3 (ALA) with PhenoAge had the greatest effect size (*B* (95% CI) = −0.69 years (−1.31, 0.06)).

**Figure 3:**
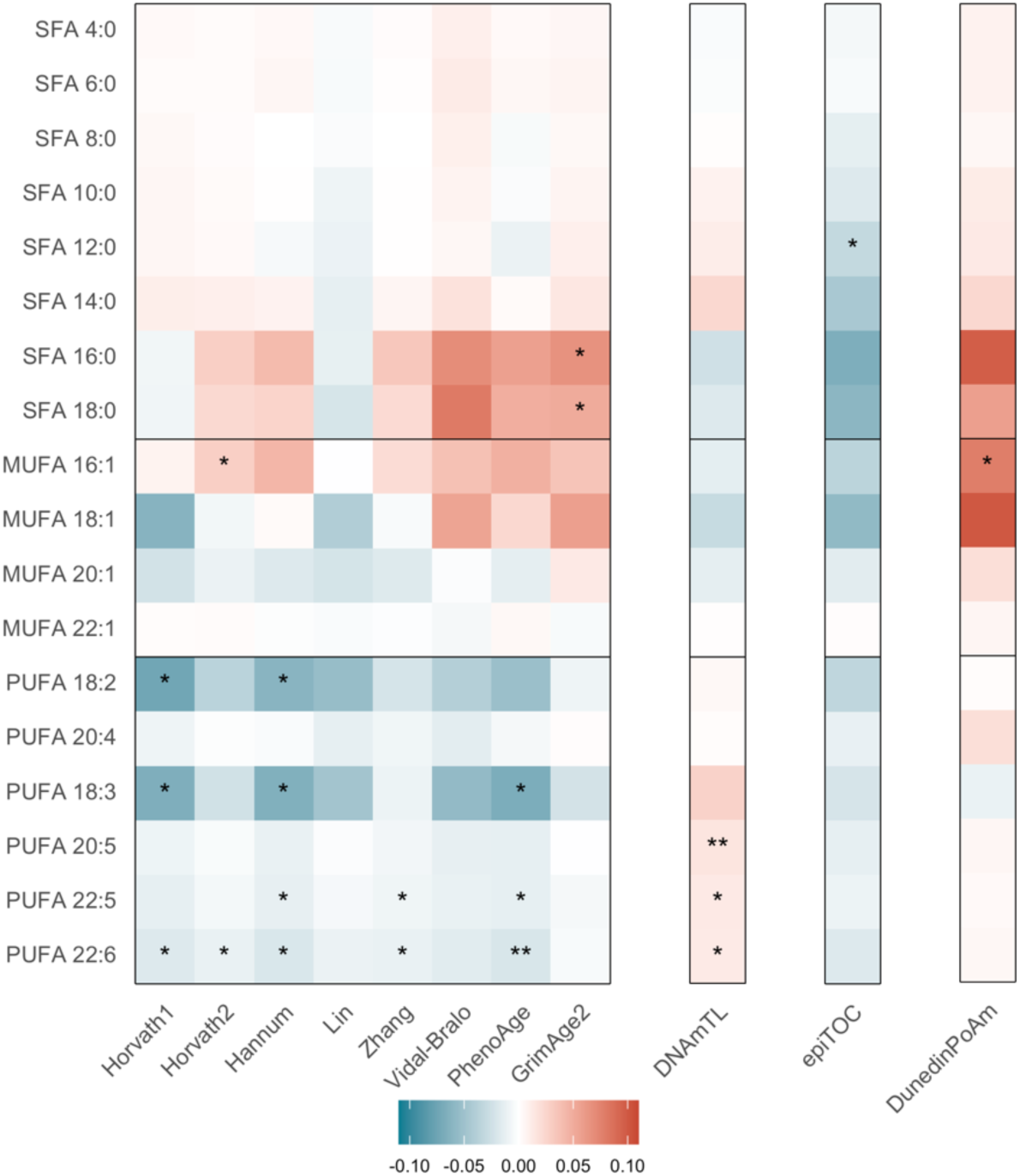
Adjusted associations of fatty acid subtype intake with epigenetic aging biomarkers. Results are from weighted generalized linear regression models adjusted for age, age^2^, sex, race and ethnicity, BMI, education level, occupation, poverty to income ratio, smoking status, alcohol intake, physical activity, and total energy intake. Effect estimates per standard deviation increase in epigenetic aging biomarkers for a log_2_ increase, or doubling, of each fatty acid subtype are represented by tile shading. * *p* < 0.05; ** *p* < 0.01.

Two SFA subtypes (SFA 16:0 and SFA 18:0) had significant positive associations with GrimAge2 (*B* (95% CI) = 0.55 years (0.07, 1.04) and 0.43 years (0.06, 0.80), respectively). The directions of association of MUFA subtypes with epigenetic aging biomarkers varied; however, MUFA 16:1 was significantly associated with greater Horvath2 and DunedinPoAm (*B* (95% CI) = 0.26 years (0.02, 0.49) and 0.08 SDs (0.01, 0.15), respectively).

### Sensitivity Analyses

We conducted sensitivity analyses using imputed covariate data (N = 2,220). The directions of association and effect sizes were largely consistent with those from analyses of complete cases (**Supplemental Table S6**). Associations of SFA and MUFA intake with GrimAge2 remained positive but imprecise (*B* (95% CI) = 0.39 years (−0.03, 0.82) and 0.40 years (−0.05, 0.86), respectively). Associations of omega-6 intake with Horvath1 and Hannum remained negative but were not statistically significant (*B* (95% CI) = −0.53 years (−1.08, 0.02) and −0.46 years (−0.98, 0.06), respectively), as did the association of omega-3 intake with Vidal-Bralo (*B* (95% CI) = −0.47 years (−0.98, 0.04)) and associations of the P:S ratio with Hannum and Vidal-Bralo (*B* (95% CI) = −0.72 years (−1.46, 0.01) and −0.70 years (−1.41, 0.02), respectively). However, with imputed covariate data, higher omega-3 intake was significantly associated with lower Horvath2 (*B* (95% CI) = −0.37 (−0.73, −0.02) and greater DNAmTL (B (95% CI) = 0.02 (0.00, 0.03)). Similarly, overall, analyses of associations with fatty acid subtypes using imputed covariate data were consistent with our primary analyses (**Supplemental Tables S7-S9**).

## Discussion

Using a nationally representative sample of US adults older than 50, we investigated the associations of fatty acid intake with epigenetic aging biomarkers, which are measures of biological aging. This study builds on previous research linking diet quality, macronutrients, and micronutrients with epigenetic aging.^14–17^ Across multiple epigenetic clocks, we found that higher intake of total PUFAs, omega-6, and omega-3, and the P:S ratio was associated with lower epigenetic aging after adjusting for BMI, total energy intake, demographics, and behavioral and lifestyle factors. Conversely, higher intake of SFAs and MUFAs was associated with increased GrimAge2, a strong biomarker of mortality risk.

Furthermore, analyses of fatty acid subtypes showed distinct, though imprecise, patterns of association: PUFA subtypes were negatively associated with epigenetic aging, SFA subtypes were positively associated with epigenetic aging, and associations of MUFA subtypes varied by aging biomarker. Overall, associations with DNAmTL, epiTOC, and DunedinPoAm were null; however, we did observe several significant associations of fatty acid subtypes with these biomarkers.

The most consistent and robust associations were observed for PUFAs. Although omega-3 fatty acids constitute a small proportion of total PUFA intake (mean omega-3 = 1.4 g/day; mean PUFA = 13.0 g/day in our sample), these fatty acids appeared to drive many of the observed associations. Higher omega-3 intake was associated with significantly lower epigenetic aging as measured by the Horvath1, Hannum, Vidal-Bralo, and PhenoAge clocks. We also found imprecise but negative associations with the Horvath2, Lin, Zhang, and GrimAge2 clocks, and a positive association with DNAmTL. Although it is difficult to distinguish between the effects of total PUFA, omega-6, and omega-3 intake due to the high correlation of these variables, the hypothesis that omega-3 fatty acids have the greatest contribution to associations with lower epigenetic aging is supported by results from models jointly including both omega-6 and omega-3 intake, as well as analyses of fatty acid subtypes. In jointly adjusted models, omega-3 intake was significantly associated with lower Vidal-Bralo and PhenoAge, whereas associations of omega-6 intake were null. In addition, three omega-3 subtypes (PUFA 18:3 (ALA), PUFA 22:5 (DPA), and PUFA 22:6 (DHA)) were significantly and negatively associated with PhenoAge; association of omega-6 subtypes with PhenoAge were not statistically significant.

Omega-3 intake may act to slow age-related decline through pathways including regulation of lipid metabolism and inflammation via activation of peroxisome proliferator-activated receptors (PPAR),^18^ regulation of immune responses such as cytokine production,^19^ and maintenance of DHA in brain cell membranes.^20^ There has been particular interest in the effects of omega-3 fatty acids in reducing cardiometabolic risks. Higher omega-3 intake has been associated with decreased triglyceride levels^21^ and lower risk of heart failure,^22^ coronary heart disease,^23^ myocardial infarction,^24^ ischemic events,^25^ and all-cause mortality;^26^ although other trials and meta-analyses have reported null associations with cardiovascular diseases^27,28^ or effects limited to omega-3 fatty acids from marine sources.^26^ However, our results and previous studies support the hypothesis of health-promoting effects of omega-3 intake through impacts on biological aging. In NHANES (1999-2018), higher omega-3 intake was associated with lower Phenotypic Age acceleration, a composite blood-based biomarker.^29^ Self-reported omega-3 intake has also been associated with lower GrimAge1 in the Framingham Heart Study.^10^ Moreover, in a recent randomized controlled trial among older adults of 1 g/day omega-3 supplementation, 2,000 IU/day vitamin D, and/or exercise (N = 777), participants receiving omega-3 had significantly lower epigenetic aging as measured by PhenoAge, GrimAge2, and DunedinPACE compared to the placebo group.^30^ The greatest effect was observed among participants receiving all three interventions and lower PhenoAge.

Considering the opposing associations of SFAs and PUFAs with the regulation of LDL cholesterol levels and chronic disease risks, we also tested for associations of the P:S ratio with epigenetic aging. A higher P:S ratio was significantly associated with lower epigenetic aging as measured by Horvath1, Hannum, Vidal-Bralo, and PhenoAge; associations with Horvath2, Zhang, GrimAge2, and DunedinPoAm were negative but imprecise. Notably, a one-unit increase in the P:S ratio, which approximates the IQR, was associated with 1.05 years lower PhenoAge. These results suggest that the P:S ratio may be a more informative aging indicator of disease-related fatty acid status than SFAs or PUFAs alone. Higher P:S ratios have previously been associated with lower LDL cholesterol.^31^ The P:S ratio may also contribute to age-related disease risk through its effect on prandial, or post-meal, levels of blood lipids. A cross-over study of meals with varying P:S ratios found a greater increase in LDL cholesterol following low P:S ratio meals compared to meals with a P:S ratio ≥ 1.^32^

Although we found only two statistically significant associations of MUFA subtypes with epigenetic aging biomarkers (MUFA 16:1 associated with greater Horvath2 and DunedinPoAm), there were notable trends in associations across MUFA subtypes. Most associations of MUFA 16:1 indicated increased epigenetic aging, associations of MUFA 18:1 were varied, and associations of MUFA 20:1 indicated decreased epigenetic aging. This heterogeneity in associations may be due to variation in food sources and prevalence in the US diet. MUFA 16:1 (hexadecenoic acid), obtained from animal and plant food sources, and MUFA 20:1 (eicosenoic acid), found in small amounts in plant oils, constituted a small proportion of total MUFA intake in our sample (mean = 1.19 and 0.18 g/day, respectively). However, the majority of MUFA intake was in the form of MUFA 18:1 (octadecenoic acid; mean = 24.0 g/day). MUFA 18:1 is common in olive and other vegetable oils, with the top food sources among NHANES participants (2005-2006) being desserts, chicken dishes, processed meats, and nuts/seeds.^33^

Overall, total fatty acid, SFA, and MUFA intake were not significantly associated with epigenetic aging. However, we did find significant positive associations of SFA and MUFA intake with GrimAge2. Among epigenetic aging biomarkers, GrimAge2 may be particularly sensitive to fatty acids with obesogenic or proinflammatory properties. GrimAge2 is calculated as a composite of sex, age, and DNA methylation surrogates for smoking pack years and nine plasma proteins, including markers of appetite regulation (leptin), insulin sensitivity (hemoglobin A1C), and inflammation (C-reactive protein (CRP), beta-2 microglobulin (B2M), and growth differentiation factor 15 (GDF15)). Feeding trials in humans have found that diets high in saturated fat are associated with increased expression of genes involved in inflammatory pathways^34^ and insulin resistance.^35^ Unexpectedly, with the exception of MUFA 16:1, we did not observe significant associations of fatty acid intake with DunedinPoAm. DunedinPoAm was trained on longitudinal changes in 18 biomarkers from adults aged 26-38, including lipoprotein(a), triglycerides, total cholesterol, and HDL cholesterol.^36^ Although DunedinPoAm has been associated with physical and cognitive decline and overall mortality, it may be less sensitive to variation in individual nutritional factors, cross-sectional associations, and in older populations.

Overall, our study included eleven epigenetic aging biomarkers available for NHANES: six first-generation clocks, four second-generation clocks (including predictors of telomere length and cell divisions), and the pace of aging. We found that PUFA and PUFA subtype intake and the P:S ratio were associated with lower epigenetic aging as measured by the Horvath1^37^ and Hannum^38^ first-generation clocks, seminal epigenetic clocks that have been associated with a broad range of health outcomes and dietary, behavioral, and environmental factors.^39,40^ However, we also identified significant associations of PUFA subtypes or the P:S ratio with the newer, and less widely used, Zhang^41^ and Vidal-Bralo^42^ clocks. The Zhang clock relied on a large training set (> 13,000 samples) to increase precision^41^ and demonstrated the greatest correlation with chronological age in our NHANES sample (*r* = 0.91). To our knowledge, and as summarized in a recent systematic review, the Zhang clock has not previously been associated with dietary, lifestyle, or behavioral factors.^39^ The Vidal-Bralo clock, which is based on eight CpGs, was developed for use with low-cost DNA methylation assays,^42^ and found to be independent of smoking status.^42,43^ Although this clock has previously been associated with physical activity in NHANES,^44^ its application has been limited. Our results demonstrate that a diversity of epigenetic clocks may be sensitive to modifiable factors such as diet.

Our study was strengthened by the use of a large, nationally representative sample of US adults. We were able to analyze the relationship of fatty acid subtypes with multiple epigenetic aging biomarkers. Consistency of associations across these measures supports the robustness of our findings. However, our study was also limited by its cross-sectional design. Longitudinal analyses are needed to evaluate the association of long-term fatty acid intake, changes in epigenetic aging, and downstream health outcomes.

We were also limited by estimates of fatty acid intake based on a 1-day dietary recall, which may not fully capture long-term dietary patterns and is vulnerable to misreporting and recall error. Fatty acid intake estimates also excluded supplement use, which may result in underestimates particularly for omega-3 intake. Furthermore, epigenetic aging biomarker data are only available in NHANES for participants aged ≥ 50 years of age in the 1999-2002 cycles. Therefore, we were not able to evaluate the relationship between fatty acid intake and epigenetic aging in younger populations or more recent NHANES cycles, which may have different dietary patterns. Generalizability may also be impacted by limitations of epigenetic clocks, which may produce biased estimates when applied to diverse populations.^45^ In addition, covariate data were missing for a portion of our sample. While our primary analyses used complete cases, analyses using imputed data were largely consistent. Finally, significance was evaluated at a nominal *p*-value < 0.05. We did not correct for multiple testing and acknowledge that some of our findings may be due to chance. The use of nominal *p*-values was due in part to our focus on overall trends in associations, and in part to the use of survey weighting, which decreases statistical power by inflating the variance of parameter estimates.^46^ Interpretation of our results, therefore, should focus on the precision of effect sizes.

### Conclusions

Fatty acids are an important dietary component involved in pathways related to both disease development and healthy aging. In this study, we provide evidence that fatty acid intake is associated with epigenetic aging biomarkers. We observed the strongest and most robust associations of higher intake of PUFAs and the P:S ratio with lower epigenetic aging across multiple clocks. These findings contribute to the growing body of research linking dietary factors to biomarkers of longevity and may inform dietary interventions and the potential utility of epigenetic clocks in clinical settings. Randomized clinical trials are needed to establish causality in the context of various dietary patterns. In addition, future studies with long-term follow-up should focus on downstream health outcomes associated with changes in epigenetic aging.

## Methods

### Study population

Analyses were conducted using data obtained from the NHANES database for the 1999-2000 and 2001-2002 survey cycles. NHANES is conducted by the US National Center for Health Statistics (NCHS) to evaluate and monitor health and nutrition among noninstitutionalized individuals in the US, and includes interviews, physical exams, and laboratory measurements. Study protocols were approved by the NCHS Research Ethics Review Board, and all participants provided written informed consent.

Epigenetic age estimates were available for 2,532 NHANES participants. The ages of participants older than 84 years were top coded as 85 to maintain confidentiality. Participants aged ≥ 85 years old (n = 130) were excluded from our analyses to prevent bias from underestimation of chronological age for older adults. We excluded participants with a mismatch between recorded sex and a DNA methylation-based sex estimate (n = 56). We further excluded 69 participants missing data dietary questionnaire data, 17 participants with unreliable dietary data, and 40 participants with extreme total energy intake (< 500 or > 5,000 kcal/day for female participants [n = 27] and < 500 or > 8,000 kcal/day for male participants [n = 13]). Our primary analyses included 1,771 participants with complete epigenetic age, fatty intake, and covariate data, and sensitivity analyses included 2,220 with complete epigenetic age and fatty intake data (**Supplemental Figure S1**).

### Fatty acid intake

Total nutrient intakes are included in the NHANES database and were estimated from 24-hour dietary recall questionnaires. Methods and data can be accessed on the NHANES website at https://www.n.cdc.gov/nchs/nhanes/search/datapage.aspx?Component=Dietary&CycleBeginYear=1999 and https://www.n.cdc.gov/nchs/nhanes/search/datapage.aspx?Component=Dietary&CycleBeginYear=2001.

In 1999-2001, the questionnaire was implemented using the NHANES computer-assisted dietary interview system (CADI). In 2002, the questionnaire was implemented as part of an integration of NHANES and the US Department of Agriculture’s (USDA’s) Continuing Survey of Food Intakes by Individuals (CSFI) using the USDA Automated Multiple Pass Method (AMPM). The 2002 data collection included a 2-day dietary recall; only the day-1 recall was used to estimate intakes released by NHANES. Total energy and nutrient intake per day were calculated by NHANES based on USD codes assigned to individual foods. This study used data on estimated daily intake in g of total fatty acids, saturated fatty acids (SFA), monounsaturated fatty acids (MUFA), polyunsaturated fatty acids (PUFA), and fatty acid subtypes, including SFA 4:0 (butanoic acid), SFA 6:0 (hexanoic acid), SFA 8:0 (octanoic acid), SFA 10:0 (decanoic acid), SFA 12:0 (dodecanoic acid), SFA 14:0 (tetradecanoic acid), SFA 16:0 (hexadecenoic acid), SFA 18:0 (octadecanoic acid), MUFA 16:1 (hexadecenoic acid), MUFA 18:1 (octadecenoic acid), MUFA 20:1 (eicosenoic acid), MUFA 22:1 (docosenoic acid), PUFA 18:2 (octadecadienoic acid, also known as linoleic acid (LA)), PUFA 18:3 (octadecatrienoic acid, also known as alpha-linolenic acid (ALA)), PUFA 18:4 (octadecatetraenoic acid, also known as stearidonic acid (SDA)), PUFA 20:4 (eicosatetraenoic acid (ETA)), PUFA 20:5 (eicosapentaenoic acid (EPA)), PUFA 22:5 (docosapentaenoic acid (DPA)), PUFA 22:6 (docosahexaenoic acid (DHA)). Total omega-6 fatty acid intake was calculated as the sum of PUFA 18:2 and PUFA 20:4, and total omega-3 fatty acid intake was calculated as the sum of PUFA 18:3, PUFA 18:4, PUFA 20:5, PUFA 22:5, and PUFA 22:6.^47,48^ The PUFA:SFA (P:S) ratio was calculated by dividing PUFA by SFA.

### DNA methylation

DNA methylation was measured in a subset of adults from the 1999-2000 and 2001-2002 survey cycles. Laboratory methods and data processing are described in detail on the NHANES website at https://www.n.cdc.gov/Nchs/Nhanes/DNAm/Default.aspx. Briefly, DNA was extracted from whole blood and stored at −80°C until DNA methylation measurement. Leukocyte DNA methylation was measured in samples from 2,532 participants aged ≥ 50 years of age with available biospecimens. Approximately half of all eligible non-Hispanic White participants were randomly selected, as well as all eligible participants from other racial or ethnic groups. DNA was bisulfite-converted and DNA methylation was measured using the Illumina Infinium MethylationEPIC BeadChip v1.0 (Illumina, San Diego, CA, US) according to the manufacturer’s instructions. Data preprocessing and quality control were performed in R and included color correction, background subtraction, removal of outlier samples based on median intensity values, and normalization with the beta mixture quantile (BMIQ) method.^49^ The calculation of the Horvath1, Horvath2, Hannum, GrimAge2, and PhenoAge biomarkers (described below) a modified version of BMIQ was used to normalize data to gold standard reference.^37^

### Epigenetic age and biomarkers

The NHANES database includes aging biomarkers that were calculated from DNA methylation data. Among available biomarkers, we limited our analyses to those that were trained on microarray data: Horvath1 (pan-tissue),^37^ Horvath2 (skin & blood),^50^ Hannum,^38^ Lin,^51^ Zhang,^41^ Vidal-Bralo,^42^ PhenoAge,^9^ GrimAge2,^11^ DNA methylation telomere length (DNAmTL),^52^ epigenetic Timer Of Cancer (epiTOC),^53^ and Dunedin Pace of Aging Methylation (DunedinPoAm).^36^ GrimAge2 is an updated version of the GrimAge mortality risk biomarker. GrimAge and GrimAge2 are both available in NHANES and are highly correlated (*r* = 0.99); therefore, our analyses only included GrimAge2.

### Covariates

Demographic and socioeconomic data, anthropometric measures, and health-related behaviors were collected as part of the NHANES questionnaires and physical examination. Methods and data can be accessed at https://www.n.cdc.gov/nchs/nhanes/continuousnhanes/default.aspx?BeginYear=1999 and https://www.n.cdc.gov/nchs/nhanes/continuousnhanes/default.aspx?BeginYear=2001. Chronological age was calculated from self-reported date of birth. Body mass index (BMI) was calculated in kg/m^2^ from height and weight collected by trained technicians. Participant race and ethnicity, as defined by NHANES, were classified as Non-Hispanic White, Mexican American, other Hispanic, Non-Hispanic Black, or other race, including Multiracial based on questions about national origin, ancestry, and self-identity. Smoking status was classified as never (having smoked < 100 cigarettes in one’s lifetime), former (having smoked ≥ 100 cigarettes in one’s lifetime but not currently smoking), or current (having smoked ≥ 100 cigarettes in one’s lifetime and currently smoking every day or some days). Alcohol consumption was based on the average number of drinks per day. Physical activity was categorized as reporting having performed moderate or vigorous activities for at least 10 minutes in the past 30 days, compared to not performing or unable to do physical activity. Occupational status was classified as white-collar and professional, white-collar and semi-routine, blue-collar and high-skill, blue-collar and semi-routine, or no work as described.^54^ Education was classified as < high school, high school diploma or GED, or > high school. The poverty to family income ratio (PIR) was calculated as the family’s income divided by the poverty guidelines for family size and year based on the Department of Health and Human Services (HHS) guidelines.

### Statistical analyses

We calculated unweighted descriptive statistics using means and standard deviations (SDs) for continuous variables and frequencies and proportions for categorical variables. We assessed performance of epigenetic clocks using Persons’s correlations and median absolute errors (MAEs) to compare chronological age and estimated epigenetic age. The survey design was specified using the *Survey* R package^55,56^ with survey weights provided with the NHANES epigenetic biomarkers dataset. To preserve the study design, the survey design was specified prior to dropping participants ≥ 85 years old, with sex mismatches, or with missing or unreliable dietary data. Correlations between fatty acid variables were calculated using the *svycor* function using the bootstrap procedure in the *jtools* R package.^57^

We evaluated associations of total fatty acids, SFA, MUFA, PUFA, omega-6, and omega-3, fatty acid subtypes, and the P:S ratio with epigenetic aging biomarkers using survey-design weighted generalized linear regression models. PUFA 18:4 was excluded from analyses of individual fatty acid subtypes due to low levels of intake. Prior to analysis, total fatty acids, SFA, MUFA, PUFA, omega-6, omega-3, and fatty acid subtypes in grams were log_2_ transformed, and for variables containing values of 0, 0.0001 was added prior to transformation. Analyses were adjusted for chronological age, chronological age^2^, sex, race and ethnicity, BMI, education level, occupation, poverty to income ratio, smoking status, alcohol intake, physical activity, and total energy intake. Including chronological age as a covariate is similar to analyzing associations with epigenetic age acceleration, also known as epigenetic age deviation, calculated as the residuals of regressing epigenetic age on chronological age. We additional conducted analyses simultaneously modeling omega-6 and omega-3 intake and tested for multicollinearity by calculating the VIF using the function *svyvif* for survey designs.^58^

Sensitivity analyses were conducted following imputation of missing covariate data. Data were imputed with multiple imputation by chained equations using the *MICE* R package^59^ and 5 iterations. Following imputation, we reanalyzed associations of fatty acid variables with aging biomarkers using fully adjusted weighted generalized linear regression models and pooled estimates from each imputed dataset with the *pool* function.

We used 95% confidence intervals (95% CIs) to evaluate precision of associations and *p* < 0.05 to test for statistical significance. All analyses were conducted in R version 4.4.1.^60^

## Data availability

All data sets analyzed in the current study are publicly available from the NHANES website (https://www.n.cdc.gov/nchs/nhanes).

## Code availability

Code for analyses and figures is available at https://github.com/annebozack/NHANES_epigeneticAge_fattyAcids.

## Acknowledgements

AKB is supported by the National Institutes of Health (NIH) grant K99ES035109. JNE and AC are supported by the NIH grant R01ES031259. This research was also supported by the National Institute on Minority Health and Health Disparities (R01MD011721, MPI: BLN and DHR; and R01MD016595). MGF is supported by the Novo Nordisk Foundation (NNF) grant NNF23SA0084103. The funders had no role in the decision to publish, preparation, review, or approval of the manuscript.

## Author contributions

AKB designed and conducted research, analyzed data, performed statistical analysis, and wrote the article; DK, JCN-E, NG, HS, SD contributed to the analyses; HS calculated the epigenetic ages; DK, JCN-E, NG, HS, SD, BLN, DHR, MG-F, and AC reviewed and edited the manuscript; AC supervised the work; AKB has primary responsibility for final content; and all authors: read and approved the final manuscript.

