## Supplemental Material for "Dietary Fatty Acids and Epigenetic Aging in US Adults: Results from the National Health and Nutrition Examination Survey"

**Figure S1: Participant flowchart.**

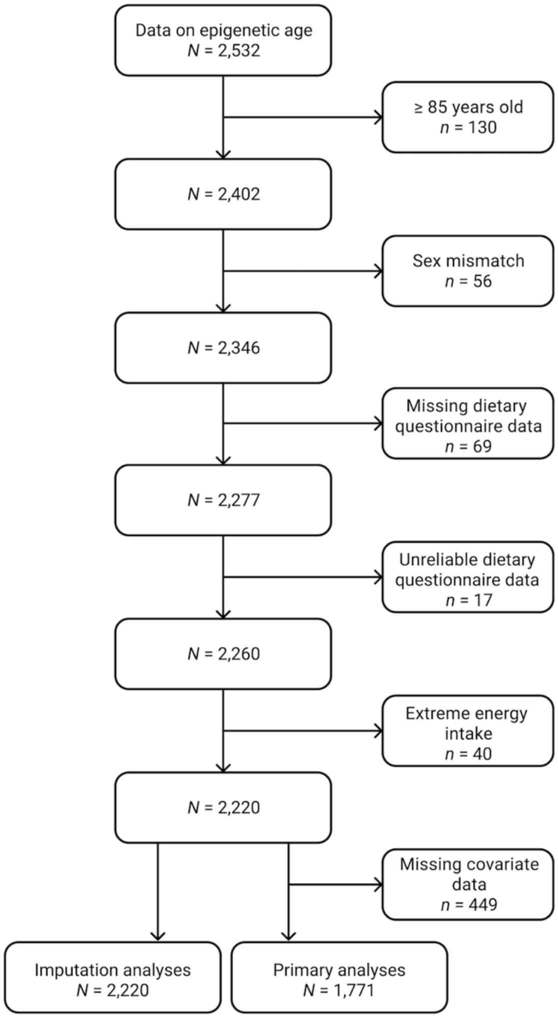

**Table S1: Characteristics of participants included in imputation analyses (N = 2,220).**

| | **Mean (SD) or n (%)** |
|---|---|
| Male, n (%) | 1,141 (51.4%) |
| Age, mean (SD) | 65.0 (9.3) |
| Race and ethnicity, n (%) | |
| Mexican American | 645 (29.2%) |
| Other Hispanic | 143 (6.4%) |
| Non-Hispanic White | 889 (40.0%) |
| Non-Hispanic Black | 467 (21.0%) |
| Other including Multiracial | 73 (3.3%) |
| BMI, mean (SD) | 28.8 (5.8) |
| Missing, n (%) | 61 (2.7%) |
| Smoking status, n (%) | |
| Never | 1,004 (45.2%) |
| Former | 863 (38.9%) |
| Current | 349 (15.7%) |
| Missing, n (%) | 2 (0.2%) |
| Alcohol intake, drinks/day, mean (SD) | 0.4 (0.9) |
| Missing, n (%) | 70 (3.2%) |
| Education, n (%) | |
| Less Than High School | 995 (44.8%) |
| More Than High School | 770 (34.7%) |
| High School Diploma (including GED) | 455 (20.5%) |
| Occupation, n (%) | |
| Blue-collar and semi-routine | 857 (38.6%) |
| Blue-collar and high-skill | 301 (13.6%) |
| White-collar and semi-routine | 376 (16.9%) |
| White-collar and professional | 505 (22.7%) |
| No work as described | 55 (2.5%) |
| Missing, n (%) | 126 (5.7%) |
| PIR, mean (SD) | 2.6 (1.6) |
| Missing, n (%) | 246 (11.1%) |
| Energy (kcal), mean (SD) | 1,830 (771) |
| Total fatty acids (g), mean (SD) | 68.1 (37.5) |
| Percent of total energy, mean (SD) | 32.8 (9.3) |
| Saturated fatty acids (g), mean (SD) | 21.5 (13.3) |
| Percent of total energy, mean (SD) | 10.3 (3.8) |
| Monounsaturated fatty acids (g), mean (SD) | 25.3 (15.1) |
| Percent of total energy, mean (SD) | 12.1 (4.1) |
| Polyunsaturated fatty acids (g), mean (SD) | 14.5 (9.5) |
| Percent of total energy, mean (SD) | 7.0 (3.3) |
| Omega-6 (g), mean (SD) | 13.0 (8.6) |
| Omega-3 (g), mean (SD) | 1.4 (1.1) |
| Polyunsaturated:saturated fatty acid ratio, mean (SD) | 0.79 (0.47) |

**Table S2: Intake of fatty acid subtypes.**

| | Primary analyses (N = 1,771) | Imputation analyses (N = 2,220) |
|---|---|---|
| **SFA 4:0 (Butanoic) (g), mean (SD)** | 0.40 (0.46) | 0.40 (0.45) |
| Intake of 0 g | 147 (8.3%) | 190 (8.6%) |
| **SFA 6:0 (Hexanoic) (g), mean (SD)** | 0.22 (0.25) | 0.22 (0.25) |
| Intake of 0 g | 186 (10.5%) | 246 (11.1%) |
| **SFA 8:0 (Octanoic) (g), mean (SD)** | 0.18 (0.21) | 0.1 (0.240) |
| Intake of 0 g | 120 (6.8%) | 167 (7.5%) |
| **SFA 10:0 (Decanoic) (g), mean (SD)** | 0.34 (0.34) | 0.33 (0.35) |
| Intake of 0 g | 37 (2.1%) | 57 (2.6%) |
| **SFA 12:0 (Dodecanoic) (g), mean (SD)** | 0.61 (0.89) | 0.61 (0.90) |
| Intake of 0 g | 5 (0.3%) | 9 (0.4%) |
| **SFA 14:0 (Tetradecanoic) (g), mean (SD)** | 1.76 (1.53) | 1.73 (1.52) |
| Intake of 0 g | 0 (0.0%) | 0 (0.0%) |
| **SFA 16:0 (Hexadecanoic) (g), mean (SD)** | 12.1 (7.02) | 11.9 (7.02) |
| Intake of 0 g | 0 (0.0%) | 0 (0.0%) |
| **SFA 18:0 (Octadecanoic) (g), mean (SD)** | 5.63 (3.48) | 5.52 (3.49) |
| Intake of 0 g | 0 (0.0%) | 1 (0.0%) |
| **MUFA 16:1 (Hexadecenoic) (g), mean (SD)** | 1.19 (0.86) | 1.17 (0.85) |
| Intake of 0 g | 0 (0.0%) | 2 (0.1%) |
| **MUFA 18:1 (Octadecenoic) (g), mean (SD)** | 24.0 (14.1) | 23.5 (14.2) |
| Intake of 0 g | 0 (0.0%) | 0 (0.0%) |
| **MUFA 20:1 (Eicosenoic) (g), mean (SD)** | 0.18 (0.39) | 0.18 (0.36) |
| Intake of 0 g | 28 (1.6%) | 40 (1.8%) |
| **MUFA 22:1 (Docosenoic) (g), mean (SD)** | 0.039 (0.14) | 0.040 (0.14) |
| Intake of 0 g | 514 (29.0%) | 683 (30.8%) |
| **PUFA 18:2 (Octadecadienoic) (g), mean (SD)** | 12.9 (8.38) | 12.8 (8.57) |
| Intake of 0 g | 0 (0.0%) | 0 (0.0%) |
| **PUFA 20:4 (Eicosatetraenoic) (g), mean (SD)** | 0.14 (0.12) | 0.13 (0.12) |
| Intake of 0 g | 63 (3.6%) | 93 (4.2%) |
| **PUFA 18:3 (Octadecatrienoic) (g), mean (SD)** | 1.30 (0.89) | 1.29 (0.92) |
| Intake of 0 g | 0 (0.0%) | 0 (0.0%) |
| **PUFA 18:4 (Octadecatetraenoic) (g), mean (SD)** | 0.0064 (0.029) | 0.0069 (0.036) |
| Intake of 0 g | 1,480 (83.6%) | 1,864 (84.0%) |
| **PUFA 20:5 (Eicosapentaenoic) (g), mean (SD)** | 0.042 (0.13) | 0.043 (0.15) |
| Intake of 0 g | 675 (38.1%) | 881 (39.7%) |
| **PUFA 22:5 (Docosapentaenoic) (g), mean (SD)** | 0.018 (0.048) | 0.018 (0.047) |
| Intake of 0 g | 916 (51.7%) | 1,167 (52.6%) |
| **PUFA 22:6 (Docosahexaenoic) (g), mean (SD)** | 0.082 (0.20) | 0.083 (0.21) |
| Intake of 0 g | 406 (22.9%) | 538 (24.2%) |

**Figure S2: Survey weighted correlations of fatty acid intake (N = 1,771).** For all correlations, $p < 0.01$.

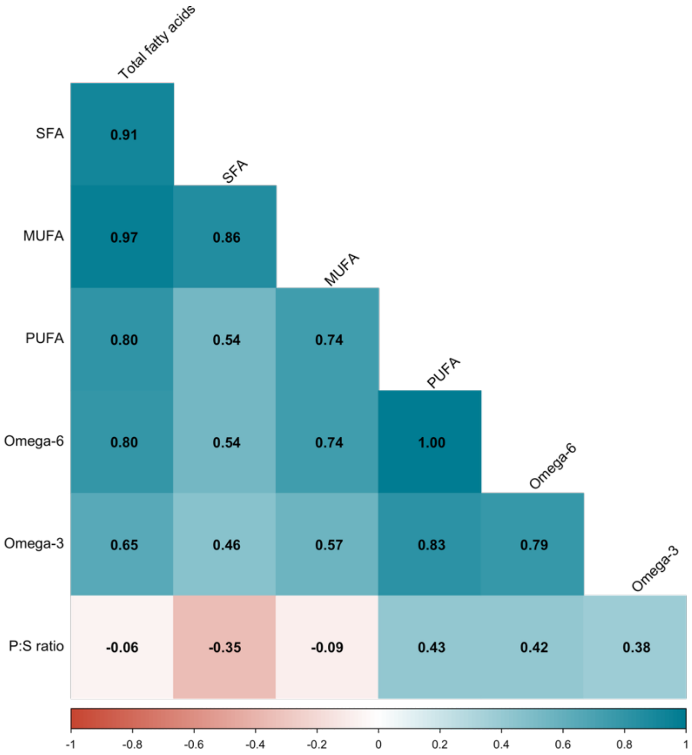

**Figure S3: Survey weighted correlations of fatty acid subtype intake (N = 1,771).** Correlations with $p < 0.05$ are shown.

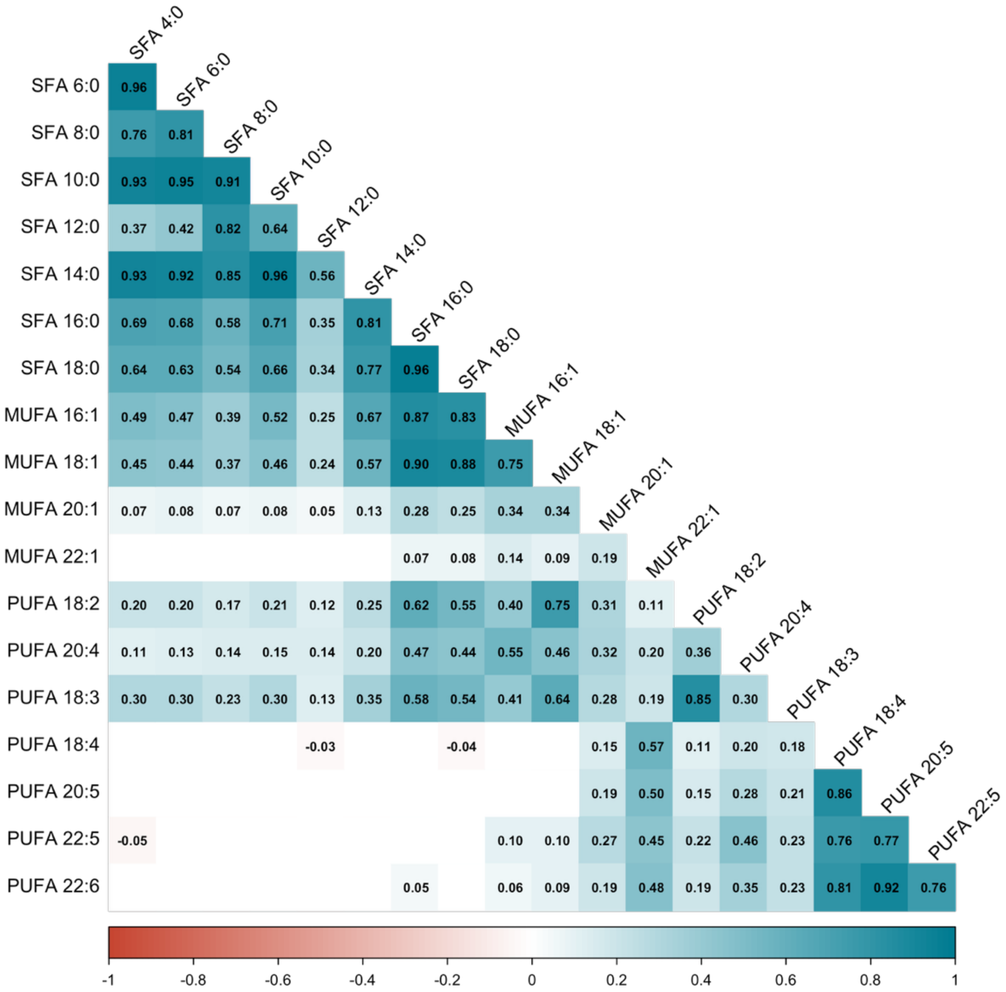

**Figure S4: Performance of epigenetic aging biomarkers (N = 1,771).** The identity line is plotted as a solid black line for the Horvath1, Horvath2, Hannum, Lin, Vidal-Bralo, Zhang, PhenoAge, and GrimAge2 clocks. MAE = median absolute error. Correlation *p*-values were < 0.001, except for DunedinPoAm ($p = 0.009$).

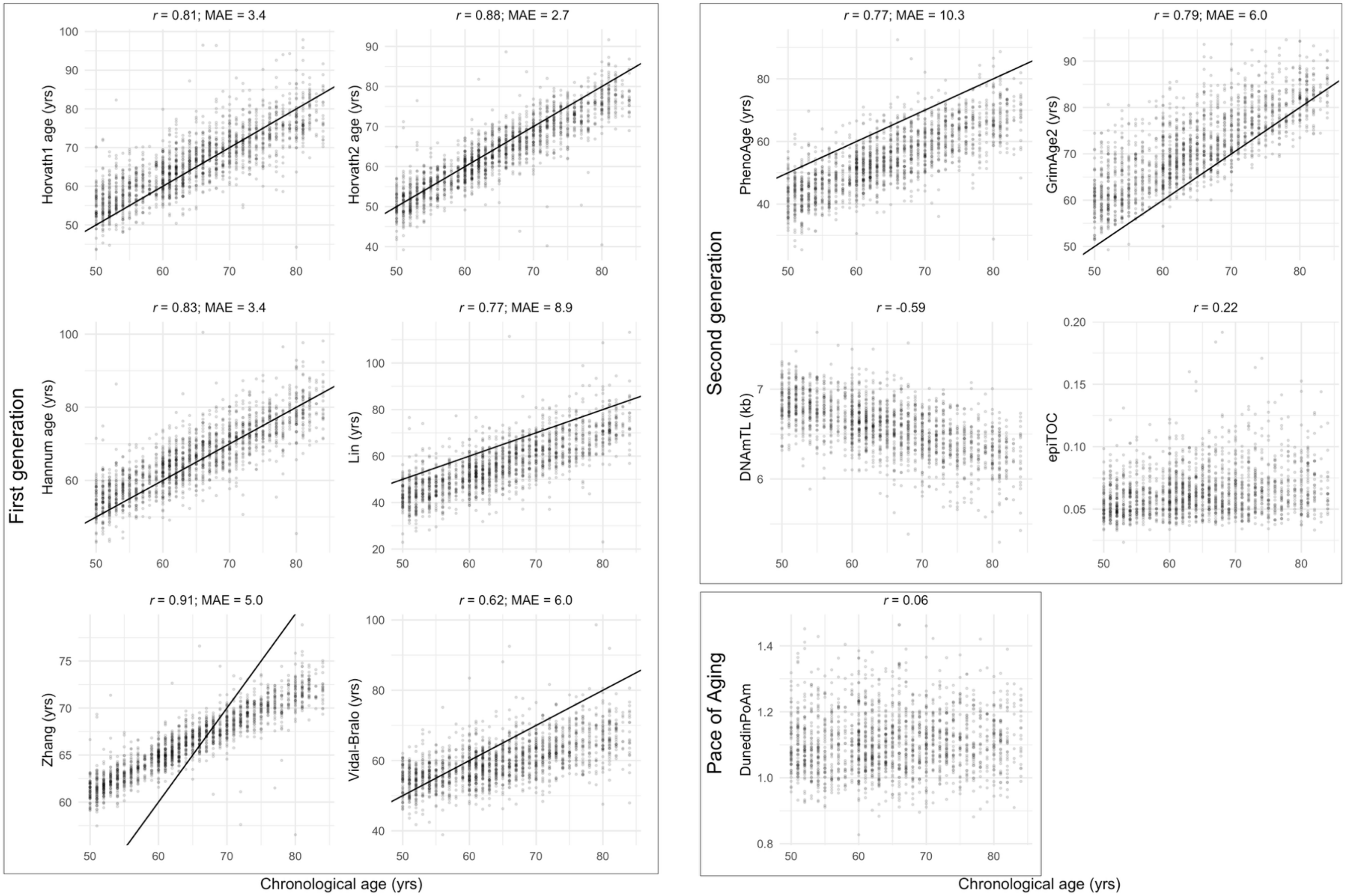

**Table S2: Associations of fatty acids and the ratio of polyunsaturated:saturated fatty acids (P:S) with epigenetic aging biomarkers (*N* = 1,771).** Effect estimates (95% confidence intervals (CIs)) are shown for a $\log_2$ increase, or doubling, of total, saturated (SFA), monounsaturated (MUFA), and polyunsaturated (PUFA) fatty acid intake, and for a one-unit increase in the P:S ratio, which approximates the interquartile range. Units are in years for the Horvath1, Horvath2, Hannum, Lin, Zhang, Vidal-Bralo, PhenoAge, and GrimAge2 clocks; kilobases for DNAmTL, and standard deviations for epiTOC and DunedinPoAm. Results are from weighted generalized linear regression models adjusted for age, $age^2$, sex, race and ethnicity, BMI, education level, occupation, poverty to income ratio, smoking status, alcohol intake, physical activity, and total energy intake.

| | **Total fatty acids** | | **SFA** | | **MFA** | | **PFA** | | **Omega-6** | | **Omega-3** | | **P:S ratio** | |
|---|---|---|---|---|---|---|---|---|---|---|---|---|---|---|
| | *B* (95% CI) | *p* | *B* (95% CI) | *p* | *B* (95% CI) | | *B* (95% CI) | *p* | *B* (95% CI) | *p* | *B* (95% CI) | *p* | *B* (95% CI) | *p* |
| Horvath1 | -0.59 (-1.40, 0.21) | 0.13 | -0.01 (-0.46, 0.45) | 0.97 | -0.50 (-1.20, 0.20) | 0.14 | -0.68 (-1.26, -0.09) | 0.029 | -0.64 (-1.23, -0.06) | 0.036 | -0.63 (-1.10, -0.16) | 0.014 | -0.87 (-1.61, -0.13) | 0.027 |
| Horvath2 | -0.07 (-0.69, 0.56) | 0.81 | 0.21 (-0.17, 0.59) | 0.23 | -0.03 (-0.54, 0.48) | 0.89 | -0.34 (-0.82, 0.14) | 0.14 | -0.33 (-0.81, 0.15) | 0.16 | -0.27 (-0.58, 0.05) | 0.09 | -0.54 (-1.19, 0.11) | 0.09 |
| Hannum | -0.20 (-1.08, 0.67) | 0.61 | 0.23 (-0.34, 0.79) | 0.38 | 0.08 (-0.72, 0.89) | 0.82 | -0.60 (-1.15, -0.06) | 0.034 | -0.57 (-1.10, -0.03) | 0.040 | -0.67 (-1.20, -0.14) | 0.020 | -0.89 (-1.62, -0.16) | 0.023 |
| Lin | -0.61 (-1.86, 0.65) | 0.30 | -0.24 (-1.09, 0.61) | 0.53 | -0.44 (-1.53, 0.66) | 0.39 | -0.64 (-1.33, 0.05) | 0.07 | -0.63 (-1.32, 0.05) | 0.07 | -0.58 (-1.19, 0.03) | 0.06 | -0.42 (-1.34, 0.49) | 0.32 |
| Zhang | 0.00 (-0.26, 0.25) | 0.97 | 0.08 (-0.07, 0.22) | 0.25 | -0.01 (-0.23, 0.21) | 0.93 | -0.09 (-0.26, 0.08) | 0.28 | -0.08 (-0.25, 0.09) | 0.32 | -0.07 (-0.20, 0.06) | 0.25 | -0.16 (-0.36, 0.04) | 0.11 |
| Vidal-Bralo | 0.19 (-0.78, 1.15) | 0.67 | 0.38 (-0.30, 1.05) | 0.23 | 0.42 (-0.44, 1.28) | 0.29 | -0.27 (-0.81, 0.27) | 0.28 | -0.25 (-0.78, 0.27) | 0.30 | -0.45 (-0.88, -0.02) | 0.041 | -0.80 (-1.56, -0.04) | 0.041 |
| PhenoAge | -0.11 (-1.20, 0.98) | 0.82 | 0.34 (-0.44, 1.13) | 0.34 | 0.32 (-0.63, 1.26) | 0.46 | -0.59 (-1.14, -0.03) | 0.040 | -0.54 (-1.08, 0.01) | 0.05 | -0.77 (-1.33, -0.21) | 0.013 | -1.05 (-1.87, -0.22) | 0.019 |
| GrimAge2 | 0.50 (-0.10, 1.09) | 0.09 | 0.42 (0.02, 0.83) | 0.043 | 0.54 (0.04, 1.04) | 0.039 | -0.08 (-0.46, 0.30) | 0.62 | -0.07 (-0.45, 0.30) | 0.66 | -0.19 (-0.60, 0.21) | 0.30 | -0.49 (-1.04, 0.06) | 0.07 |
| DNAmTL | 0.00 (-0.02, 0.03) | 0.99 | 0.00 (-0.02, 0.02) | 0.99 | -0.01 (-0.03, 0.01) | 0.32 | 0.00 (-0.01, 0.02) | 0.71 | 0.00 (-0.02, 0.02) | 0.85 | 0.01 (0.00, 0.03) | 0.12 | 0.00 (-0.02, 0.03) | 0.68 |
| epiTOC | -0.09 (-0.22, 0.05) | 0.18 | -0.08 (-0.18, 0.02) | 0.10 | -0.06 (-0.18, 0.07) | 0.31 | -0.04 (-0.12, 0.04) | 0.30 | -0.03 (-0.11, 0.04) | 0.33 | -0.04 (-0.12, 0.04) | 0.28 | 0.03 (-0.08, 0.15) | 0.52 |
| DunedinPoAm | 0.08 (-0.06, 0.22) | 0.21 | 0.07 (-0.04, 0.18) | 0.19 | 0.11 (-0.01, 0.23) | 0.08 | 0.00 (-0.08, 0.09) | 0.90 | 0.00 (-0.09, 0.09) | 0.94 | 0.01 (-0.08, 0.10) | 0.83 | -0.05 (-0.15, 0.04) | 0.24 |

**Table S3: Associations of omega-3 and omega-6 intake with epigenetic aging biomarkers in jointly adjusted models (*N* = 1,771).** Effect estimates (95% confidence intervals (CIs)) are shown for a $\log_2$ increase, or doubling, of total, saturated (SFA), monounsaturated (MUFA), and polyunsaturated (PUFA) fatty acid intake, and for a one-unit increase in the P:S ratio. Units are in years for the Horvath1, Horvath2, Hannum, Lin, Zhang, Vidal-Bralo, PhenoAge, and GrimAge2 clocks; kilobases for DNAmTL, and standard deviations for epiTOC and DunedinPoAm. Results are from weighted generalized linear regression models including both omega-6 and omega-3 intake adjusted for age, $age^2$, sex, race and ethnicity, BMI, education level, occupation, poverty to income ratio, smoking status, alcohol intake, physical activity, and total energy intake.

| | **Omega-6** | | **Omega-3** | |
|---|---|---|---|---|
| | *B* (95% CI) | *p* | *B* (95% CI) | *p* |
| Horvath1 | -0.31 (-1.24, 0.62) | 0.46 | -0.44 (-1.17, 0.30) | 0.21 |
| Horvath2 | -0.23 (-0.97, 0.50) | 0.48 | -0.12 (-0.63, 0.39) | 0.59 |
| Hannum | -0.10 (-0.91, 0.71) | 0.78 | -0.61 (-1.39, 0.18) | 0.11 |
| Lin | -0.36 (-1.46, 0.75) | 0.47 | -0.36 (-1.34, 0.63) | 0.42 |
| Zhang | -0.05 (-0.28, 0.18) | 0.65 | -0.04 (-0.21, 0.13) | 0.59 |
| Vidal-Bralo | 0.19 (-0.53, 0.90) | 0.56 | -0.57 (-1.13, -0.01) | 0.047 |
| PhenoAge | 0.11 (-0.67, 0.89) | 0.75 | -0.84 (-1.64, -0.05) | 0.041 |
| GrimAge2 | 0.14 (-0.46, 0.75) | 0.59 | -0.28 (-0.93, 0.36) | 0.33 |
| DNAmTL | -0.02 (-0.05, 0.01) | 0.17 | 0.03 (0.00, 0.06) | 0.08 |
| epiTOC | -0.01 (-0.12, 0.10) | 0.86 | -0.03 (-0.15, 0.08) | 0.50 |
| DunedinPoAm | -0.01 (-0.14, 0.12) | 0.92 | 0.01 (-0.11, 0.14) | 0.83 |

**Table S3: Associations of saturated fatty acid (SFA) subtypes with epigenetic aging biomarkers (*N* = 1,771).** Effect estimates (95% confidence intervals (CIs)) are shown for a $\log_2$ increase, or doubling, of fatty acid intake. Units are in years for the Horvath1, Horvath2, Hannum, Lin, Zhang, Vidal-Bralo, PhenoAge, and GrimAge2 clocks; kilobases for DNAmTL, and standard deviations for epiTOC and DunedinPoAm. Results are from weighted generalized linear regression models adjusted for age, age$^2$, sex, race and ethnicity, BMI, education level, occupation, poverty to income ratio, smoking status, alcohol intake, physical activity, and total energy intake.

| | **SFA 4:0** | | **SFA 6:0** | | **SFA 8:0** | | **SFA 10:0** | | **SFA 12:0** | | **SFA 14:0** | | **SFA 16:0** | | **SFA 18:0** | |
|---|---|---|---|---|---|---|---|---|---|---|---|---|---|---|---|---|
| | *B* (95% CI) | *p* | *B* (95% CI) | *p* | *B* (95% CI) | | *B* (95% CI) | *p* | *B* (95% CI) | *p* | *B* (95% CI) | *p* | *B* (95% CI) | *p* | *B* (95% CI) | *p* |
| Horvath1 | 0.03 (-0.05, 0.11) | 0.43 | 0.02 (-0.08, 0.12) | 0.65 | 0.04 (-0.07, 0.15) | 0.47 | 0.05 (-0.10, 0.19) | 0.49 | 0.05 (-0.10, 0.20) | 0.49 | 0.09 (-0.13, 0.31) | 0.38 | -0.06 (-0.63, 0.51) | 0.81 | -0.07 (-0.54, 0.41) | 0.75 |
| Horvath2 | 0.02 (-0.04, 0.08) | 0.40 | 0.02 (-0.04, 0.09) | 0.45 | 0.02 (-0.07, 0.11) | 0.65 | 0.03 (-0.10, 0.15) | 0.63 | 0.03 (-0.12, 0.18) | 0.63 | 0.09 (-0.15, 0.32) | 0.42 | 0.26 (-0.14, 0.66) | 0.18 | 0.21 (-0.10, 0.51) | 0.16 |
| Hannum | 0.04 (-0.07, 0.14) | 0.45 | 0.05 (-0.05, 0.15) | 0.31 | 0.00 (-0.13, 0.12) | 0.97 | 0.00 (-0.16, 0.16) | 0.96 | -0.04 (-0.23, 0.14) | 0.60 | 0.07 (-0.21, 0.35) | 0.59 | 0.38 (-0.30, 1.06) | 0.23 | 0.24 (-0.30, 0.77) | 0.33 |
| Lin | -0.04 (-0.25, 0.16) | 0.62 | -0.05 (-0.21, 0.12) | 0.53 | -0.03 (-0.27, 0.2) | 0.76 | -0.10 (-0.37, 0.16) | 0.39 | -0.12 (-0.45, 0.22) | 0.44 | -0.15 (-0.59, 0.29) | 0.46 | -0.14 (-1.17, 0.88) | 0.76 | -0.25 (-0.97, 0.47) | 0.45 |
| Zhang | 0.01 (-0.02, 0.03) | 0.55 | 0.00 (-0.03, 0.04) | 0.78 | 0.00 (-0.04, 0.04) | 0.99 | 0.00 (-0.04, 0.05) | 0.91 | 0.00 (-0.05, 0.05) | 0.94 | 0.02 (-0.06, 0.10) | 0.56 | 0.12 (-0.04, 0.28) | 0.13 | 0.08 (-0.05, 0.21) | 0.21 |
| Vidal-Bralo | 0.06 (-0.07, 0.19) | 0.32 | 0.08 (-0.04, 0.19) | 0.16 | 0.06 (-0.08, 0.20) | 0.35 | 0.04 (-0.12, 0.21) | 0.56 | 0.03 (-0.20, 0.26) | 0.76 | 0.11 (-0.22, 0.44) | 0.46 | 0.45 (-0.32, 1.21) | 0.22 | 0.52 (-0.17, 1.21) | 0.12 |
| PhenoAge | 0.03 (-0.12, 0.19) | 0.62 | 0.05 (-0.09, 0.19) | 0.44 | -0.04 (-0.19, 0.10) | 0.50 | -0.03 (-0.2, 0.14) | 0.71 | -0.10 (-0.31, 0.10) | 0.28 | 0.03 (-0.28, 0.34) | 0.84 | 0.58 (-0.36, 1.53) | 0.19 | 0.49 (-0.29, 1.27) | 0.18 |
| GrimAge2 | 0.04 (-0.09, 0.18) | 0.48 | 0.06 (-0.07, 0.18) | 0.33 | 0.04 (-0.08, 0.16) | 0.50 | 0.06 (-0.09, 0.20) | 0.39 | 0.08 (-0.08, 0.24) | 0.29 | 0.12 (-0.10, 0.34) | 0.24 | 0.55 (0.07, 1.04) | 0.031 | 0.43 (0.06, 0.80) | 0.029 |
| DNAmTL | 0.00 (0.00, 0.00) | 0.47 | 0.00 (0.00, 0.00) | 0.54 | 0.00 (0.00, 0.01) | 0.84 | 0.00 (0.00, 0.01) | 0.42 | 0.00 (0.00, 0.01) | 0.36 | 0.01 (0.00, 0.02) | 0.16 | -0.01 (-0.03, 0.01) | 0.41 | -0.01 (-0.02, 0.01) | 0.50 |
| epiTOC | -0.01 (-0.02, 0.01) | 0.43 | 0.00 (-0.02, 0.01) | 0.58 | -0.01 (-0.03, 0.01) | 0.15 | -0.02 (-0.04, 0.01) | 0.11 | -0.030 (-0.06, 0.00) | 0.045 | -0.04 (-0.09, 0.00) | 0.06 | -0.07 (-0.18, 0.05) | 0.22 | -0.06 (-0.15, 0.03) | 0.17 |
| DunedinPoAm | 0.01 (-0.01, 0.02) | 0.30 | 0.01 (-0.01, 0.03) | 0.33 | 0.00 (-0.02, 0.03) | 0.63 | 0.01 (-0.02, 0.04) | 0.41 | 0.01 (-0.03, 0.05) | 0.50 | 0.02 (-0.03, 0.08) | 0.38 | 0.10 (-0.03, 0.22) | 0.12 | 0.06 (-0.05, 0.16) | 0.24 |

**Table S4: Associations of monounsaturated fatty acid (MUFA) subtypes with epigenetic aging biomarkers (*N* = 1,771).** Effect estimates (95% confidence intervals (CIs)) are shown for a $\log_2$ increase, or doubling, of fatty acid intake. Units are in standard deviations for epigenetic aging biomarkers. Results are from weighted generalized linear regression models adjusted for age, $age^2$, sex, race and ethnicity, BMI, education level, occupation, poverty to income ratio, smoking status, alcohol intake, physical activity, and total energy intake.

| | **MUFA 16:1** | | **MUFA 18:1** | | **MUFA 20:1** | | **MUFA 22:1** | |
|---|---|---|---|---|---|---|---|---|
| | *B* (95% CI) | *p* | *B* (95% CI) | *p* | *B* (95% CI) | | *B* (95% CI) | *p* |
| Horvath1 | 0.06 (-0.28, 0.40) | 0.68 | -0.53 (-1.22, 0.15) | 0.11 | -0.20 (-0.55, 0.15) | 0.22 | 0.02 (-0.11, 0.14) | 0.79 |
| Horvath2 | 0.26 (0.02, 0.49) | 0.034 | -0.06 (-0.58, 0.46) | 0.79 | -0.09 (-0.33, 0.14) | 0.39 | 0.02 (-0.07, 0.11) | 0.58 |
| Hannum | 0.40 (-0.04, 0.84) | 0.07 | 0.03 (-0.78, 0.83) | 0.94 | -0.16 (-0.52, 0.20) | 0.32 | -0.02 (-0.13, 0.10) | 0.76 |
| Lin | -0.01 (-0.59, 0.58) | 0.98 | -0.47 (-1.53, 0.58) | 0.33 | -0.26 (-0.59, 0.07) | 0.11 | -0.03 (-0.17, 0.10) | 0.58 |
| Zhang | 0.07 (-0.02, 0.17) | 0.11 | -0.01 (-0.23, 0.20) | 0.89 | -0.06 (-0.17, 0.05) | 0.26 | 0.00 (-0.04, 0.03) | 0.76 |
| Vidal-Bralo | 0.24 (-0.19, 0.68) | 0.24 | 0.36 (-0.47, 1.18) | 0.35 | -0.01 (-0.25, 0.23) | 0.90 | -0.03 (-0.12, 0.05) | 0.36 |
| PhenoAge | 0.48 (-0.04, 1.01) | 0.07 | 0.24 (-0.68, 1.16) | 0.57 | -0.14 (-0.47, 0.19) | 0.36 | 0.04 (-0.10, 0.19) | 0.50 |
| GrimAge2 | 0.30 (-0.1, 0.70) | 0.12 | 0.49 (-0.01, 0.99) | 0.05 | 0.11 (-0.05, 0.27) | 0.15 | -0.03 (-0.11, 0.04) | 0.33 |
| DNAmTL | 0.00 (-0.02, 0.01) | 0.52 | -0.01 (-0.03, 0.01) | 0.38 | 0.00 (-0.02, 0.01) | 0.42 | 0.00 (0.00, 0.00) | 0.86 |
| epiTOC | -0.04 (-0.11, 0.04) | 0.30 | -0.06 (-0.18, 0.07) | 0.31 | -0.02 (-0.06, 0.03) | 0.48 | 0.00 (-0.01, 0.02) | 0.83 |
| DunedinPoAm | 0.08 (0.01, 0.15) | 0.036 | 0.10 (-0.02, 0.22) | 0.10 | 0.02 (-0.02, 0.05) | 0.24 | 0.01 (-0.01, 0.02) | 0.44 |

**Table S5: Associations of polyunsaturated fatty acid (PUFA) subtypes with epigenetic aging biomarkers (*N* = 1,771).** Effect estimates (95% confidence intervals (CIs)) are shown for a $\log_2$ increase, or doubling, of fatty acid intake. Units are in standard deviations for epigenetic aging biomarkers. Results are from weighted generalized linear regression models adjusted for age, $age^2$, sex, race and ethnicity, BMI, education level, occupation, poverty to income ratio, smoking status, alcohol intake, physical activity, and total energy intake.

| | **PUFA 18:2** | | **PUFA 20:4** | | **PUFA 18:3** | | **PUFA 20:5** | | **PUFA 22:5** | | **PUFA 22:6** | |
|---|---|---|---|---|---|---|---|---|---|---|---|---|
| | *B* (95% CI) | *p* | *B* (95% CI) | *p* | *B* (95% CI) | | *B* (95% CI) | *p* | *B* (95% CI) | *p* | *B* (95% CI) | *p* |
| Horvath1 | -0.63 (-1.22, -0.05) | 0.037 | -0.08 (-0.23, 0.07) | 0.28 | -0.57 (-1.04, -0.10) | 0.024 | -0.08 (-0.20, 0.04) | 0.17 | -0.11 (-0.22, 0.00) | 0.06 | -0.15 (-0.27, -0.03) | 0.018 |
| Horvath2 | -0.32 (-0.80, 0.15) | 0.16 | -0.01 (-0.12, 0.10) | 0.78 | -0.22 (-0.56, 0.12) | 0.17 | -0.03 (-0.11, 0.05) | 0.40 | -0.05 (-0.12, 0.01) | 0.10 | -0.10 (-0.19, -0.02) | 0.018 |
| Hannum | -0.56 (-1.09, -0.03) | 0.040 | -0.03 (-0.2, 0.15) | 0.72 | -0.60 (-1.13, -0.06) | 0.033 | -0.10 (-0.21, 0.00) | 0.06 | -0.12 (-0.22, -0.01) | 0.033 | -0.18 (-0.31, -0.05) | 0.011 |
| Lin | -0.63 (-1.30, 0.05) | 0.07 | -0.15 (-0.35, 0.05) | 0.12 | -0.56 (-1.17, 0.06) | 0.07 | -0.02 (-0.16, 0.11) | 0.68 | -0.06 (-0.20, 0.08) | 0.33 | -0.12 (-0.27, 0.03) | 0.10 |
| Zhang | -0.08 (-0.24, 0.09) | 0.32 | -0.03 (-0.07, 0.02) | 0.23 | -0.03 (-0.17, 0.11) | 0.59 | -0.02 (-0.05, 0.01) | 0.11 | -0.03 (-0.06, 0.00) | 0.036 | -0.04 (-0.08, 0.00) | 0.038 |
| Vidal-Bralo | -0.25 (-0.77, 0.27) | 0.30 | -0.10 (-0.23, 0.04) | 0.14 | -0.36 (-0.82, 0.10) | 0.11 | -0.08 (-0.18, 0.02) | 0.11 | -0.07 (-0.16, 0.02) | 0.09 | -0.10 (-0.2, 0.00) | 0.05 |
| PhenoAge | -0.53 (-1.07, 0.01) | 0.05 | -0.06 (-0.26, 0.15) | 0.55 | -0.69 (-1.31, -0.06) | 0.036 | -0.13 (-0.26, 0.00) | 0.05 | -0.13 (-0.26, 0.00) | 0.049 | -0.21 (-0.34, -0.07) | 0.008 |
| GrimAge2 | -0.08 (-0.45, 0.30) | 0.65 | 0.01 (-0.09, 0.12) | 0.79 | -0.19 (-0.62, 0.23) | 0.32 | 0.00 (-0.06, 0.05) | 0.89 | -0.05 (-0.12, 0.03) | 0.18 | -0.03 (-0.09, 0.02) | 0.20 |
| DNAmTL | 0.00 (-0.02, 0.02) | 0.86 | 0.00 (0.00, 0.01) | 0.82 | 0.01 (-0.01, 0.03) | 0.38 | 0.00 (0.00, 0.01) | 0.007 | 0.00 (0.00, 0.01) | 0.026 | 0.00 (0.00, 0.01) | 0.032 |
| epiTOC | -0.03 (-0.11, 0.04) | 0.35 | -0.01 (-0.04, 0.01) | 0.33 | -0.02 (-0.11, 0.06) | 0.57 | -0.01 (-0.03, 0.00) | 0.07 | -0.01 (-0.03, 0.01) | 0.20 | -0.02 (-0.04, 0.00) | 0.06 |
| DunedinPoAm | 0.00 (-0.09, 0.09) | 0.96 | 0.02 (-0.01, 0.05) | 0.19 | -0.01 (-0.10, 0.08) | 0.78 | 0.01 (-0.01, 0.02) | 0.48 | 0.00 (-0.02, 0.02) | 0.68 | 0.00 (-0.01, 0.02) | 0.59 |

**Table S6: Associations of fatty acid intake with epigenetic aging biomarkers using imputed covariate data (*N* = 2,220).** Effect estimates (95% confidence intervals (CIs)) are shown for a $\log_2$ increase, or doubing, of total, saturated (SFA), monounsaturated (MFA), and polyunsaturated (PFA), omega-6, and omega-3 fatty acids or for a one-unit increase in polyunsaturated:saturated fatty acid (P:S) ratio, which approximates the interquartile range. Results are from weighted generalized linear regression models adjusted for age, $age^2$, sex, race and ethnicity, BMI, education level, occupation, poverty to income ratio, smoking status, alcohol intake, physical activity, and total energy intake.

| | **Total fatty acids** | | **SFA** | | **MFA** | | **PFA** | | **Omega-6** | | **Omega-3** | | **P:S ratio** | |
|---|---|---|---|---|---|---|---|---|---|---|---|---|---|---|
| | *B* (95% CI) | *p* | *B* (95% CI) | *p* | *B* (95% CI) | | *B* (95% CI) | *p* | *B* (95% CI) | *p* | *B* (95% CI) | *p* | *B* (95% CI) | *p* |
| Horvath1 | -0.50 (-1.23, 0.23) | 0.15 | 0.04 (-0.39, 0.46) | 0.85 | -0.42 (-1.07, 0.22) | 0.16 | -0.60 (-1.13, -0.08) | 0.029 | -0.53 (-1.08, 0.02) | 0.05 | -0.61 (-1.07, -0.14) | 0.019 | -0.83 (-1.45, -0.21) | 0.016 |
| Horvath2 | -0.19 (-0.74, 0.37) | 0.45 | 0.10 (-0.33, 0.52) | 0.60 | -0.12 (-0.58, 0.34) | 0.56 | -0.37 (-0.78, 0.03) | 0.06 | -0.34 (-0.76, 0.09) | 0.10 | -0.37 (-0.73, -0.02) | 0.043 | -0.49 (-1.11, 0.13) | 0.10 |
| Hannum | -0.23 (-1.06, 0.59) | 0.52 | 0.13 (-0.42, 0.68) | 0.58 | 0.00 (-0.75, 0.75) | 1.00 | -0.53 (-1.04, -0.02) | 0.043 | -0.46 (-0.98, 0.06) | 0.08 | -0.66 (-1.17, -0.14) | 0.020 | -0.72 (-1.46, 0.01) | 0.05 |
| Lin | -0.48 (-1.68, 0.73) | 0.38 | -0.33 (-1.04, 0.38) | 0.30 | -0.25 (-1.30, 0.79) | 0.58 | -0.46 (-1.15, 0.23) | 0.16 | -0.39 (-1.12, 0.35) | 0.25 | -0.52 (-1.19, 0.16) | 0.11 | -0.23 (-1.06, 0.61) | 0.54 |
| Zhang | -0.04 (-0.26, 0.18) | 0.66 | 0.03 (-0.11, 0.17) | 0.61 | -0.03 (-0.23, 0.16) | 0.71 | -0.09 (-0.23, 0.05) | 0.15 | -0.08 (-0.22, 0.06) | 0.23 | -0.12 (-0.24, 0.01) | 0.07 | -0.14 (-0.30, 0.03) | 0.09 |
| Vidal-Bralo | 0.11 (-0.93, 1.16) | 0.80 | 0.33 (-0.34, 1.00) | 0.28 | 0.23 (-0.67, 1.12) | 0.56 | -0.26 (-0.84, 0.31) | 0.31 | -0.20 (-0.78, 0.37) | 0.42 | -0.47 (-0.98, 0.04) | 0.06 | -0.70 (-1.41, 0.02) | 0.05 |
| PhenoAge | -0.22 (-1.21, 0.76) | 0.61 | 0.23 (-0.45, 0.91) | 0.44 | 0.15 (-0.75, 1.04) | 0.71 | -0.57 (-1.12, -0.01) | 0.047 | -0.46 (-1.05, 0.13) | 0.11 | -0.85 (-1.48, -0.22) | 0.016 | -0.97 (-1.81, -0.13) | 0.029 |
| GrimAge2 | 0.36 (-0.18, 0.90) | 0.16 | 0.39 (-0.03, 0.82) | 0.06 | 0.40 (-0.05, 0.86) | 0.07 | -0.16 (-0.53, 0.21) | 0.33 | -0.11 (-0.48, 0.26) | 0.49 | -0.33 (-0.71, 0.04) | 0.07 | -0.55 (-1.12, 0.02) | 0.06 |
| DNAmTL | 0.01 (-0.02, 0.03) | 0.62 | 0.01 (-0.01, 0.02) | 0.51 | -0.01 (-0.03, 0.02) | 0.48 | 0.00 (-0.01, 0.02) | 0.57 | 0.00 (-0.01, 0.02) | 0.67 | 0.02 (0.00, 0.04) | 0.033 | 0.01 (-0.02, 0.03) | 0.68 |
| epiTOC | -0.06 (-0.19, 0.07) | 0.30 | -0.07 (-0.16, 0.02) | 0.11 | -0.04 (-0.16, 0.08) | 0.46 | -0.02 (-0.10, 0.06) | 0.53 | -0.02 (-0.09, 0.06) | 0.59 | -0.03 (-0.10, 0.05) | 0.41 | 0.04 (-0.07, 0.14) | 0.42 |
| DunedinPoAm | 0.05 (-0.09, 0.18) | 0.42 | 0.05 (-0.05, 0.15) | 0.27 | 0.08 (-0.02, 0.18) | 0.10 | -0.02 (-0.10, 0.07) | 0.64 | -0.01 (-0.10, 0.07) | 0.74 | -0.01 (-0.10, 0.07) | 0.70 | -0.08 (-0.17, 0.02) | 0.10 |

**Table S7: Associations of saturated fatty acid (SFA) subtypes with epigenetic aging biomarkers using imputed covariate data (*N* = 2,220).** Effect estimates (95% confidence intervals (CIs)) are shown for a $\log_2$ increase, or doubling, of fatty acid intake. Units are in years for the Horvath1, Horvath2, Hannum, Lin, Zhang, Vidal-Bralo, PhenoAge, and GrimAge2 clocks; kilobases for DNAmTL, and standard deviations for epiTOC and DunedinPoAm. Results are from weighted generalized linear regression models adjusted for age, $age^2$, sex, race and ethnicity, BMI, education level, occupation, poverty to income ratio, smoking status, alcohol intake, physical activity, and total energy intake.

| | **SFA 4:0** | | **SFA 6:0** | | **SFA 8:0** | | **SFA 10:0** | | **SFA 12:0** | | **SFA 14:0** | | **SFA 16:0** | | **SFA 18:0** | |
|---|---|---|---|---|---|---|---|---|---|---|---|---|---|---|---|---|
| | *B* (95% CI) | *p* | *B* (95% CI) | *p* | *B* (95% CI) | | *B* (95% CI) | *p* | *B* (95% CI) | *p* | *B* (95% CI) | *p* | *B* (95% CI) | *p* | *B* (95% CI) | *p* |
| Horvath1 | 0.05 (-0.02, 0.12) | 0.13 | 0.03 (-0.07, 0.13) | 0.45 | 0.03 (-0.09, 0.15) | 0.54 | 0.05 (-0.09, 0.20) | 0.39 | 0.04 (-0.11, 0.19) | 0.57 | 0.12 (-0.10, 0.33) | 0.23 | -0.01 (-0.52, 0.49) | 0.95 | -0.03 (-0.51, 0.46) | 0.90 |
| Horvath2 | 0.03 (-0.04, 0.10) | 0.30 | 0.01 (-0.06, 0.09) | 0.69 | 0.00 (-0.12, 0.11) | 0.92 | 0.00 (-0.13, 0.13) | 0.98 | -0.01 (-0.17, 0.15) | 0.85 | 0.08 (-0.17, 0.34) | 0.47 | 0.14 (-0.29, 0.58) | 0.46 | 0.10 (-0.3, 0.51) | 0.56 |
| Hannum | 0.04 (-0.05, 0.14) | 0.35 | 0.03 (-0.07, 0.13) | 0.51 | -0.03 (-0.16, 0.10) | 0.59 | -0.02 (-0.18, 0.14) | 0.79 | -0.06 (-0.26, 0.14) | 0.48 | 0.06 (-0.23, 0.36) | 0.64 | 0.28 (-0.33, 0.89) | 0.31 | 0.15 (-0.39, 0.68) | 0.54 |
| Lin | -0.02 (-0.21, 0.17) | 0.77 | -0.04 (-0.22, 0.13) | 0.56 | -0.07 (-0.3, 0.17) | 0.51 | -0.14 (-0.37, 0.09) | 0.20 | -0.20 (-0.48, 0.09) | 0.15 | -0.19 (-0.58, 0.19) | 0.27 | -0.21 (-1.07, 0.65) | 0.58 | -0.31 (-0.98, 0.35) | 0.30 |
| Zhang | 0.01 (-0.02, 0.03) | 0.48 | 0.00 (-0.04, 0.03) | 0.88 | -0.01 (-0.06, 0.03) | 0.52 | -0.01 (-0.05, 0.03) | 0.70 | -0.01 (-0.06, 0.04) | 0.57 | 0.01 (-0.06, 0.09) | 0.68 | 0.07 (-0.08, 0.22) | 0.29 | 0.03 (-0.11, 0.18) | 0.59 |
| Vidal-Bralo | 0.10 (-0.02, 0.21) | 0.09 | 0.10 (0.01, 0.18) | 0.037 | 0.08 (-0.03, 0.18) | 0.13 | 0.05 (-0.10, 0.19) | 0.47 | 0.03 (-0.16, 0.23) | 0.69 | 0.14 (-0.15, 0.43) | 0.29 | 0.38 (-0.38, 1.15) | 0.27 | 0.41 (-0.28, 1.09) | 0.20 |
| PhenoAge | 0.06 (-0.06, 0.17) | 0.29 | 0.05 (-0.06, 0.15) | 0.30 | -0.03 (-0.16, 0.09) | 0.54 | -0.04 (-0.19, 0.10) | 0.52 | -0.12 (-0.31, 0.07) | 0.18 | 0.01 (-0.26, 0.27) | 0.96 | 0.39 (-0.41, 1.20) | 0.28 | 0.38 (-0.36, 1.11) | 0.26 |
| GrimAge2 | 0.04 (-0.06, 0.15) | 0.35 | 0.05 (-0.05, 0.16) | 0.24 | 0.03 (-0.06, 0.13) | 0.43 | 0.05 (-0.07, 0.17) | 0.37 | 0.08 (-0.07, 0.23) | 0.25 | 0.11 (-0.11, 0.34) | 0.25 | 0.43 (-0.06, 0.93) | 0.07 | 0.41 (0.01, 0.8) | 0.045 |
| DNAmTL | 0.00 (0.00, 0.00) | 0.58 | 0.00 (0.00, 0.00) | 0.83 | 0.00 (0.00, 0.01) | 0.44 | 0.00 (0.00, 0.01) | 0.35 | 0.00 (0.00, 0.01) | 0.21 | 0.01 (0.00, 0.02) | 0.11 | 0.00 (-0.02, 0.02) | 0.90 | 0.00 (-0.02, 0.02) | 0.85 |
| epiTOC | 0.00 (-0.02, 0.01) | 0.42 | -0.01 (-0.02, 0.01) | 0.37 | -0.01 (-0.03, 0.00) | 0.08 | -0.02 (-0.04, 0.00) | 0.08 | -0.03 (-0.05, 0.00) | 0.043 | -0.04 (-0.08, 0.01) | 0.08 | -0.04 (-0.16, 0.07) | 0.42 | -0.05 (-0.14, 0.04) | 0.24 |
| DunedinPoAm | 0.00 (-0.01, 0.02) | 0.70 | 0.00 (-0.01, 0.02) | 0.56 | 0.00 (-0.02, 0.02) | 0.87 | 0.01 (-0.02, 0.03) | 0.59 | 0.00 (-0.03, 0.04) | 0.80 | 0.01 (-0.04, 0.07) | 0.56 | 0.06 (-0.07, 0.19) | 0.29 | 0.05 (-0.06, 0.15) | 0.33 |

**Table S8: Associations of monounsaturated fatty acid (MUFA) subtypes with epigenetic aging biomarkers using imputed covariate data (*N* = 2,220).** Effect estimates (95% confidence intervals (CIs)) are shown for a $\log_2$ increase, or doubling, of fatty acid intake. Units are in standard deviations for epigenetic aging biomarkers. Results are from weighted generalized linear regression models adjusted for age, $age^2$, sex, race and ethnicity, BMI, education level, occupation, poverty to income ratio, smoking status, alcohol intake, physical activity, and total energy intake.

| | **MUFA 16:1** | | **MUFA 18:1** | | **MUFA 20:1** | | **MUFA 22:1** | |
|---|---|---|---|---|---|---|---|---|
| | *B* (95% CI) | *p* | *B* (95% CI) | *p* | *B* (95% CI) | | *B* (95% CI) | *p* |
| Horvath1 | 0.08 (-0.17, 0.34) | 0.46 | -0.44 (-1.07, 0.20) | 0.15 | -0.09 (-0.41, 0.23) | 0.52 | 0.00 (-0.11, 0.12) | 0.93 |
| Horvath2 | 0.20 (-0.02, 0.43) | 0.07 | -0.15 (-0.61, 0.30) | 0.45 | -0.01 (-0.24, 0.21) | 0.91 | -0.01 (-0.1, 0.08) | 0.74 |
| Hannum | 0.27 (-0.05, 0.59) | 0.08 | -0.04 (-0.79, 0.71) | 0.90 | -0.07 (-0.39, 0.25) | 0.62 | -0.05 (-0.17, 0.06) | 0.31 |
| Lin | -0.02 (-0.52, 0.48) | 0.93 | -0.28 (-1.32, 0.77) | 0.55 | -0.13 (-0.46, 0.21) | 0.40 | -0.03 (-0.16, 0.09) | 0.54 |
| Zhang | 0.05 (-0.02, 0.12) | 0.14 | -0.04 (-0.23, 0.15) | 0.62 | -0.03 (-0.13, 0.07) | 0.54 | -0.02 (-0.05, 0.02) | 0.30 |
| Vidal-Bralo | 0.23 (-0.13, 0.59) | 0.18 | 0.17 (-0.70, 1.03) | 0.65 | 0.06 (-0.17, 0.29) | 0.56 | -0.03 (-0.11, 0.05) | 0.38 |
| PhenoAge | 0.28 (-0.12, 0.69) | 0.14 | 0.09 (-0.79, 0.97) | 0.81 | -0.10 (-0.4, 0.20) | 0.46 | -0.02 (-0.17, 0.13) | 0.75 |
| GrimAge2 | 0.14 (-0.15, 0.42) | 0.29 | 0.37 (-0.07, 0.82) | 0.09 | 0.08 (-0.06, 0.21) | 0.21 | -0.05 (-0.12, 0.03) | 0.16 |
| DNAmTL | 0.00 (-0.01, 0.01) | 0.86 | -0.01 (-0.03, 0.02) | 0.59 | 0.00 (-0.01, 0.01) | 0.39 | 0.00 (0.00, 0.01) | 0.35 |
| epiTOC | 0.00 (-0.08, 0.07) | 0.91 | -0.04 (-0.16, 0.08) | 0.45 | 0.00 (-0.04, 0.04) | 0.87 | 0.01 (-0.01, 0.02) | 0.38 |
| DunedinPoAm | 0.03 (-0.06, 0.12) | 0.47 | 0.08 (-0.02, 0.19) | 0.11 | 0.01 (-0.02, 0.04) | 0.50 | 0.00 (-0.02, 0.01) | 0.80 |

**Table S9: Associations of polyunsaturated fatty acid (PUFA) subtypes with epigenetic aging biomarkers using imputed covariate data (*N* = 2,220).** Effect estimates (95% confidence intervals (CIs)) are shown for a $\log_2$ increase, or doubling, of fatty acid intake. Units are in standard deviations for epigenetic aging biomarkers. Results are from weighted generalized linear regression models adjusted for age, $age^2$, sex, race and ethnicity, BMI, education level, occupation, poverty to income ratio, smoking status, alcohol intake, physical activity, and total energy intake.

| | **PUFA 18:2** | | **PUFA 20:4** | | **PUFA 18:3** | | **PUFA 20:5** | | **PUFA 22:5** | | **PUFA 22:6** | |
|---|---|---|---|---|---|---|---|---|---|---|---|---|
| | *B* (95% CI) | *p* | *B* (95% CI) | *p* | *B* (95% CI) | | *B* (95% CI) | *p* | *B* (95% CI) | *p* | *B* (95% CI) | *p* |
| Horvath1 | -0.53 (-1.07, 0.01) | 0.05 | -0.04 (-0.22, 0.15) | 0.65 | -0.6 (-1.11, -0.10) | 0.025 | -0.04 (-0.17, 0.09) | 0.50 | -0.09 (-0.20, 0.02) | 0.08 | -0.09 (-0.22, 0.04) | 0.15 |
| Horvath2 | -0.33 (-0.75, 0.09) | 0.10 | 0.01 (-0.13, 0.16) | 0.84 | -0.39 (-0.81, 0.02) | 0.06 | -0.01 (-0.11, 0.09) | 0.79 | -0.05 (-0.11, 0.02) | 0.15 | -0.06 (-0.17, 0.05) | 0.22 |
| Hannum | -0.46 (-0.98, 0.06) | 0.07 | -0.02 (-0.22, 0.18) | 0.79 | -0.64 (-1.18, -0.10) | 0.026 | -0.07 (-0.19, 0.05) | 0.21 | -0.12 (-0.22, -0.01) | 0.031 | -0.12 (-0.26, 0.03) | 0.09 |
| Lin | -0.39 (-1.11, 0.34) | 0.24 | -0.10 (-0.36, 0.15) | 0.35 | -0.54 (-1.21, 0.14) | 0.10 | 0.00 (-0.17, 0.17) | 0.96 | -0.03 (-0.17, 0.11) | 0.65 | -0.04 (-0.24, 0.15) | 0.60 |
| Zhang | -0.08 (-0.22, 0.06) | 0.22 | -0.02 (-0.09, 0.04) | 0.47 | -0.10 (-0.25, 0.04) | 0.13 | -0.01 (-0.05, 0.02) | 0.39 | -0.03 (-0.05, 0.00) | 0.07 | -0.02 (-0.07, 0.02) | 0.28 |
| Vidal-Bralo | -0.21 (-0.77, 0.36) | 0.41 | -0.05 (-0.19, 0.08) | 0.39 | -0.45 (-1.01, 0.11) | 0.10 | -0.03 (-0.13, 0.06) | 0.43 | -0.04 (-0.12, 0.04) | 0.25 | -0.05 (-0.14, 0.05) | 0.28 |
| PhenoAge | -0.46 (-1.04, 0.12) | 0.10 | -0.03 (-0.23, 0.17) | 0.73 | -0.82 (-1.51, -0.13) | 0.027 | -0.10 (-0.25, 0.05) | 0.15 | -0.11 (-0.24, 0.02) | 0.09 | -0.14 (-0.29, 0.01) | 0.06 |
| GrimAge2 | -0.11 (-0.48, 0.25) | 0.49 | -0.03 (-0.12, 0.06) | 0.42 | -0.34 (-0.72, 0.04) | 0.07 | 0.00 (-0.06, 0.05) | 0.87 | -0.06 (-0.13, 0.01) | 0.08 | -0.03 (-0.08, 0.03) | 0.26 |
| DNAmTL | 0.00 (-0.01, 0.02) | 0.68 | 0.00 (0.00, 0.01) | 0.38 | 0.01 (0.00, 0.03) | 0.10 | 0.00 (0.00, 0.01) | 0.010 | 0.00 (0.00, 0.01) | 0.009 | 0.00 (0.00, 0.01) | 0.11 |
| epiTOC | -0.02 (-0.09, 0.06) | 0.60 | 0.00 (-0.02, 0.03) | 0.69 | -0.02 (-0.10, 0.06) | 0.55 | -0.01 (-0.02, 0.00) | 0.11 | -0.01 (-0.02, 0.00) | 0.10 | -0.01 (-0.03, 0.00) | 0.14 |
| DunedinPoAm | -0.01 (-0.10, 0.07) | 0.72 | 0.00 (-0.02, 0.03) | 0.73 | -0.03 (-0.11, 0.05) | 0.42 | 0.00 (-0.01, 0.02) | 0.76 | 0.00 (-0.02, 0.02) | 0.84 | 0.00 (-0.02, 0.02) | 1.00 |
